# Within-Day Variability of SARS-CoV-2 RNA in Municipal Wastewater Influent During Periods of Varying COVID-19 Prevalence and Positivity

**DOI:** 10.1101/2021.03.16.21253652

**Authors:** Aaron Bivins, Devin North, Zhenyu Wu, Marlee Shaffer, Warish Ahmed, Kyle Bibby

## Abstract

Wastewater surveillance of severe acute respiratory syndrome coronavirus 2 (SARS-CoV-2) RNA is being used to monitor Coronavirus Disease 2019 (COVID-19) trends in communities; however, within-day variation in primary influent concentrations of SARS-CoV-2 RNA remain largely uncharacterized. In the current study, grab sampling of primary influent was performed every 2 hours over two different 24-hour periods at two wastewater treatment plants (WWTPs) in northern Indiana, USA. In primary influent, uncorrected, recovery-corrected, and pepper mild mottle virus (PMMoV)-normalized SARS-CoV-2 RNA concentrations demonstrated ordinal agreement with increasing clinical COVID-19 positivity, but not COVID-19 cases. Primary influent SARS-CoV-2 RNA concentrations exhibited greater variation than PMMoV RNA concentrations as expected for lower shedding prevalence. The bovine respiratory syncytial virus (BRSV) process control recovery efficiency was low (mean: 0.91%) and highly variable (coefficient of variation: 51% - 206%) over the four sampling events with significant differences between the two WWTPs (p <0.0001). The process control recovery was similar to the independently assessed SARS-CoV-2 RNA recovery efficiency, which was also significantly different between the two WWTPs (p <0.0001). Recovery-corrected SARS-CoV-2 RNA concentrations better reflected within-day changes in primary influent flow rate and fecal content, as indicated by PMMoV concentrations. These observations highlight the importance of assessing the process recovery efficiency, which is highly variable, using an appropriate process control. Despite large variations, both recovery-corrected and PMMoV-normalized SARS-CoV-2 RNA concentrations in primary influent demonstrate potential for monitoring COVID-19 positivity trends in WWTPs serving peri-urban and rural areas.

## INTRODUCTION

When infected with severe acute coronavirus 2 (SARS-CoV-2), the β-coronavirus which causes coronavirus disease 2019 (COVID-19), humans, both symptomatic and asymptomatic^1^, shed the virus and its RNA, in various body fluids^2–4^ including: sputum, saliva, urine, and feces. Since many of these body fluids are deposited into wastewater collection systems, wastewater-based epidemiology (WBE) has emerged as a promising technique^5^ for corroborating clinical surveillance observations, or monitoring SARS-CoV-2 infection when clinical surveillance systems are unavailable or limited^6^.

Surveillance strategies and sampling methods for WBE remain diverse, with community-level temporal trends monitored via both primary solids^7,8^ and primary influent^9^. Studies monitoring primary influent for surveillance have used grab samples^10–14^, time-based composite samples^15,16^, and flow-based composite samples^9,17^. Concentrations of SARS-CoV-2 RNA in wastewater and wastewater solids correlate with COVID-19 cases^8,17,18^ and positivity rates^19^. Attempts to use wastewater data to estimate SARS-CoV-2 infection prevalence remain limited due to large uncertainty and variation in shedding rates and viral sewershed dynamics^20,21^.

To reduce variation, normalization of SARS-CoV-2 RNA concentrations by pepper mild mottle virus (PMMoV) RNA concentration has been suggested to account for the fecal content of wastewater samples^8^. PMMoV is an elongated rod-shaped virus with a single-stranded genome^22^ that is prevalent in human feces^23,24^ due to the consumption of produce and is subsequently prevalent in wastewater globally^25^. WBE studies of wastewater solids have reported improved correlation with clinical case trends^26^ and no effect^8^ associated with PMMoV-normalization, while a study of wastewater influent found that PMMoV-normalization decreased correlation with clinical case trends^18^.

The SARS-CoV-2 RNA concentration in municipal wastewater influent is expected to exhibit temporal trends consistent with domestic sewage inputs and PMMoV influent concentration due to the fecal shedding of SARS-CoV-2 RNA by those infected. This variability then drives best sampling practices, e.g., grab versus composite samples. Studies of within-day variation in SARS-CoV-2 RNA concentrations in primary influent remain limited. A recent study comparing flow-weighted composites and grab samples found agreement between the two, but suggested avoiding sampling during and immediately following early morning low flow periods due to low concentrations from grab samples^27^. Another study hypothesized that a 10-fold increase in SARS-CoV-2 RNA concentrations in flow-weighted influent composites compared to grab samples suggested diurnal variation, but called for additional testing to confirm^28^. Additional evidence is necessary to identify best sampling practices and inform data interpretation.

The purpose of the current study was to assess the variability associated with SARS-CoV-2 RNA in primary influent at two wastewater treatment plants (WWTPs) during distinct periods of epidemic COVID-19. The effort had two primary goals. The first goal was to characterize the within-day variation in influent SARS-CoV-2 RNA concentrations, PMMoV RNA concentrations, and process control recovery efficiency. The second goal was to assess the relationships between primary influent SARS-CoV-2 RNA concentration, including normalized concentrations, and COVID-19 clinical surveillance metrics.

## MATERIALS AND METHODS

### Primary Influent Sampling Locations

The experiments described herein were conducted at two WWTPs located in two communities, identified as community A and community B, in northern Indiana, USA. Records from the Environmental Protection Agency’s (EPA) Enforcement and Compliance History Online (ECHO) system indicate the design flow for each WWTP is 20 million gallons per day (MGD) (https://echo.epa.gov/). The WWTP in community A (WWTP A) serves 56,227 residents and had an average influent flow rate of 14.09 million gallons per day (MGD) in 2020 (250 gallons per capita-day) while the WWTP in community B (WWTP B) serves 46,557 residents and had an average influent flow rate of 11.50 MGD in 2020 (247 gallons per capita-day). Despite serving fewer residents, the population density surrounding WWTP B is greater (2,995 persons per square mile) than the density surrounding WWTP A (1,881 persons per square mile). COVID-19 clinical surveillance data during the 14 days prior to each sampling period for the counties A and B were obtained from the Indiana COVID-19 Dashboard and Map (https://www.coronavirus.in.gov/2393.htm). COVID-19 clinical surveillance data at the sub-county level are not publicly available for this region.

### 24-h Sampling Experiments

A total of four 24-hour sampling experiments were conducted: (1) WWTP A from 12:00 June 18 to 12:00 June 19, 2020; (2) WWTP A from 1:30 to 23:30 December 2; (3) WWTP B from 11:00 May 7 to 9:00 May 8, 2020; (4) WWTP B from 9:00 December 1 to 7:00 December 2, 2020. During each experiment, 500 mL primary influent grab samples were collected at 2-hour intervals and immediately stored at 4°C. Samples were then transported on ice to the laboratory and again stored at 4°C until concentrated as described below within 24 hours. At WWTP A, 24-hour time-based composite samples were also collected on 18 and 19 June and 2 December. While at WWTP B, a 24-hour time-based composite sample was only prepared using the grab samples from 1 December to 2 December. The average hourly flow rates were recorded during each experiment and subsequently used to calculate average flow rates for each 2-hour interval and for the entire 24-hour experiment.

### Electronegative Membrane Adsorption and Extraction

Primary influent wastewater samples were concentrated using an electronegative membrane followed by direct extraction of the membrane as has been previously reported for concentration of viral markers from surface water^29^ and SARS-CoV-2 for WBE applications^21^. During the experiments, 100 mL of primary influent was filtered through a 0.45 µm 47 mm GN-6 Metricel hydrophilic mixed cellulose ester membrane (Pall Corporation, Port Washington, NY, USA) on a glass vacuum filtration assembly (Sigma-Aldrich, St. Louis, MO, USA).

Prior to concentration, each influent sample was seeded with a process control, bovine respiratory syncytial virus (BRSV), in the form of Inforce 3, an intranasal cattle vaccine consisting of live attenuated virus (Zoetis, Parsippany-Troy Hills, NJ, USA) at a ratio of 1 µL of Inforce 3/mL of wastewater. The spike concentration was 4.73 ± 0.09 log_10_ RNA copy number/µL as quantified by direct extraction of 500 µL aliquots of seeded wastewater. BRSV was selected as a process control because of its similarity to SARS-CoV-2 morphology: both are enveloped viruses with helical symmetry and negative sense single-stranded RNA genomes^30^.

Immediately after concentration, each membrane filter was rolled and placed into a 2 mL Garnet PowerBead Tube (Qiagen, Hilden, Germany) using aseptic technique and frozen at −80°C. Nucleic acids were extracted from each sample using the AllPrep PowerViral DNA/RNA kit (Qiagen, Hilden, Germany). Prior to extraction, 800 µL of solution PM1 (heated to 55°C) and 8 µL of β-Mecaptoethanol (MP Biomedicals, Irvine, CA, USA) were added to each thawed PowerBead tube, vortexed briefly, and homogenized on a FastPrep 24 beat beating instrument for four rounds of 20 seconds at 4.5 M/s with 30 seconds rest between each round. After bead beating, the PowerBead tubes were centrifuged at 13,000 x g for one minute and 500 µL of the resulting supernatant was transferred into a clean 2 mL microcentrifuge tube. The extraction was then completed following the Qiagen protocol. In the final step, nucleic acids were eluted in 80 µL of RNase free water (provided with the kit). The resulting eluate was centrifuged for two minutes at 13,000 x g and 60 µL of supernatant was transferred into a 2 mL DNA LoBind tube (Eppendorf, Hamburg, Germany) and stored at −80°C until assayed by reverse transcription droplet digital polymerase chain reaction (RT-ddPCR).

### Direct Extractions

In addition to the primary influent samples concentrated by the adsorption-extraction method, a paired subset of 16 samples, eight collected from WWTP A and 8 from WWTP B (December 2020 experiments), were extracted by adding 500 µL of influent directly into a Garnet PowerBead tube and extracting the nucleic acids as described above. The purpose of these direct extractions was to directly estimate the virus RNA concentration recovery efficiency by comparing the direct extraction enumerations and the adsorption-extraction enumerations.

### RT-ddPCR

RNA in sample extracts was detected and quantified by RT-ddPCR performed on the BioRad QX200 Droplet Digital PCR System with thermal cycling performed on the C1000 Touch Thermal Cycler (BioRad, Hercules, CA, USA). RNA reverse transcription and PCR amplification was performed in a single reaction using the One-Step RT-ddPCR Advanced Kit for Probes (BioRad, Hercules, CA, USA) per the manufacturer’s instructions. Each reaction was prepared as a 22 µL volume consisting of 5.25 µL of 4X reaction mix, 2.1 µL of reverse transcriptase, 1.05 µL of dithiothreitol, 6.45 µL of molecular grade water, and 4 µL of nucleic acid extract from each sample. Primer and probe sequences, concentrations, and thermal cycling conditions for each RT-ddPCR assay are summarized in Table S1. Each RT-ddPCR experiment included no-template controls, positive controls, and the pertinent negative extraction controls as described in further detail below. The RNA copy number for each RT-ddPCR reaction was estimated by manual thresholding performed in QuantaSoft Version 1.7.4 (BioRad, Hercules, CA, USA) such that the negative controls, both no-template and extraction, were negative for each assay.

To assess the extraction and RT-ddPCR efficiency, a subset of 16 concentrated influent samples (8 WWTP A; 8 WWTP B) were seeded with Hepatitis G (Hep G) Armored RNA (Asuragen, Austin, TX, USA) as a molecular process control^9,31^. For these samples, 10 µL of Hep G Armored RNA was seeded into the 500 µL supernatant resulting from membrane filter homogenization and then extracted and subjected to RT-ddPCR as described above. The starting titer of the Hep G spike (1,140 ± 152 RNA GC/µL) was determined by heat-extracting an aliquot of Hep G Armored RNA at 75°C for 3 minutes and quantifying the resulting RNA by RT-ddPCR per the manufacturer’s instructions. The extraction and RT-ddPCR recovery efficiency was estimated by comparing the quantity of Hep G RNA recovered from each sample with the starting titer.

### RT-ddPCR Assays

SARS-CoV-2 RNA was detected and quantified using the CDC N1 assay targeting the nucleocapsid gene^32^. The N1 copy number in each sample was measured in triplicate RT-ddPCR reactions using the premixed primers and probe (Table S1) from the 2019-nCoV RUO Kit (IDT, Coralville, IA, USA). Each RT-ddPCR experiment included a no-template control, two positive controls consisting of the 2019-nCoV_N_Positive Control plasmid (IDT, Coralville, IA, USA), and the relevant negative extraction control. The 95% limit of detection (95%LOD) for the N1 assay was estimated using a 1:3 dilution series of Synthetic SARS-CoV-2 RNA Control (MT 188340) (Twist Bioscience, San Francisco, CA, USA). At each step in the dilution series, 12 RT-ddPCR replicates were assayed. A cumulative Gaussian distribution was fit to the observed proportion of positive technical replicates along the dilution series, and the 95% LOD was estimated as the 95th percentile of the resulting distribution.

PMMoV RNA was quantified using an RT-ddPCR assay targeting the replicase protein gene^23,33^. The forward and reverse primers (Table S1) were synthesized by IDT (Coralville, IA, USA) while the Taqman minor-grove-binder probe was synthesized by Applied Biosystems (Foster City, CA, USA). PMMoV RNA was quantified in a single RT-ddPCR reaction for each sample with two negative controls included in each experiment.

RNA from the process control, BRSV, was detected and quantified using an assay targeting the nucleoprotein gene^34^ and adapted to RT-ddPCR format^35^. The forward and reverse primers and probe (Table S1) were synthesized by IDT (Coralville, IA, USA). BRSV RNA for each sample was measured in a single RT-ddPCR reaction with two negative controls and two positive controls consisting of extract (Qiagen AllPrep PowerViral) of Inforce 3 aliquots. RNA from the extraction and molecular control, Hep G Armored RNA, was quantified in RT-ddPCR duplicates using an RT-ddPCR assay targeting polyprotein precursor^31^ with primers and probes (Table S1) synthesized by IDT (Coralville, IA, USA). RT-ddPCR experiments were deemed satisfactory when each target was detected in the relevant positive controls and not detected in its negative controls, the BRSV process control was detected in each sample, and PMMoV was detected in each sample.

### RNA Persistence Experiments

In addition to the 24-h influent sampling experiments, a daily composite sample was collected from WWTP A on 23 June 2020, seeded with BRSV, and used to investigate the stability of RNA during storage, pasteurization, and freeze-thaw cycles. To assess persistence during storage, the composite sample was aliquoted into 50 mL centrifuge tubes. The tubes were incubated at either 4°C or 25°C with two tubes combined into a single 100 mL sample and processed every 24 hours from time zero to seven days. Persistence through pasteurization was assessed by pasteurizing two 50 mL aliquots in centrifuge tubes at 60°C for 90 minutes, with a brief vortex mix at 45 minutes, and then combining the two aliquots into a single 100 mL sample. The effect of freeze-thaw cycles was assessed by freezing 50 mL aliquots in centrifuge tubes at −80°C for 48 hours, thawing the tubes at 4°C and refreezing at −80°C for up to three cycles. After each thaw, two tubes were combined into one 100 mL sample for processing. For the persistence assessments, SARS-CoV-2, PMMoV, and BRSV RNA were concentrated and extracted at each time point using the adsorption-extraction method previously described.

Comparisons between two groups were made using Mann-Whitney tests and between multiple groups using Kruskal-Wallis tests with Dunn’s correction^36–38^. All graphing and statistical analyses associated with the described experiments were performed using GraphPad Prism Version 9.0.0 (GraphPad Software, LaJolla, CA, USA).

## RESULTS AND DISCUSSION

### Process Control, Molecular Control & Concentration Recovery Efficiency

Across all experiments, 83 primary influent samples were concentrated by adsorption-extraction and assayed for SARS-CoV-2 and PMMoV RNA: 58 from WWTP A and 25 from WWTP B (one 24-hour event did not include a composite sample). The BRSV recovery efficiency across all samples processed using adsorption-extraction ranged from 0.03 to 15% with a mean of 0.91% (95%CI: 0.53 – 1.3) (Figure S1). The observed recovery efficiency in samples from WWTP A was greater than in samples from WWTP B (*p* <0.0001); however, the coefficient of variation (CV) in samples from WWTP A was also greater (169%) than WWTP B (83%). For a subset of four samples where the solids fraction was removed prior to adsorption-extraction, the BRSV mean recovery efficiency was 25% (95%CI: 22 – 28).

For a subset of 16 primary influent samples (8 from each WWTP), the recovery efficiency of the extraction and molecular control, Hep G, ranged from 38.5% to 64.4% with a mean of 49.4% (95%CI: 47.4 – 51.5) (Figure S2). Unlike the process control, the extraction and molecular control recovery from WWTPs A and B samples was not significantly different (*p* = 0.1034). Interestingly, the mean recovery for wastewater seeded with Hep G was greater than for PCR-grade water seeded with Hep G (37%, *n* = 2). Although a statistical comparison could not be made owing to the limited sample size, this suggests the extraction kit might be more efficient for wastewater than PCR-grade water.

Paired measurements of N1 copy number per liter in directly extracted influent versus influent concentrated via the adsorption-extraction method indicate that the concentration recovery efficiency for SARS-CoV-2 RNA ranged from 0.14% to 10% with an average of 1.9% (Figure S3; 95%CI: 1.4 – 2.5). Just as for the BRSV process control, the SARS-CoV-2 RNA recovery was greater in WWTP A than WWTP B (*p* < 0.0001). PMMoV RNA concentration recovery, determined in the same manner and shown in Figure S4, ranged from 7.4% to 41.3% with an average of 19.2% (95%CI: 14.8 – 23.7) and was not statistically different between the two WWTPs (*p* = 0.0771). The SARS-CoV-2 RNA concentration recovery CV of 98% was greater than PMMoV RNA recovery CV of 43%.

A wide variety of process controls have been reported in the WBE literature, including: bovine coronavirus^9,18^, f-specific RNA phages^6^, phi 6^39^, murine hepatitis virus^8,40^, vesicular stomatitis virus^26^, porcine endemic diarrhea virus^12^, mengovirus^12^, porcine respiratory and reproductive syndrome virus and murine norovirus^41^, human coronavirus OC43^42^, human coronavirus 229E^43^, and even inactivated SARS-CoV-2^40,44^. During a methods comparison, significantly different recovery efficiencies were observed between a variety of process controls for a single method^42^. In the current study SARS-CoV-2 and PMMoV were recovered at different mean efficiencies (1.9% and 19.2%, respectively) using the adsorption-extraction method. The mean SARS-CoV-2 RNA concentration recovery (1.9%) and molecular control recovery (49%) considered in series result in an estimated mean process recovery efficiency of 0.93%. This is comparable with the mean process efficiency estimated by the BRSV control (0.91%). For the workflow described in this study, BRSV seems a reasonable process control for estimating the recovery of SARS-CoV-2 RNA, given the limitations inherent to all process controls.^45^ The mean recovery of BRSV observed during this study is much lower than the 6.6% recovery reported for electronegative membrane filtration with acidification and MgCl_2_ amendment reported during a study in Virginia^9^. However, electronegative membrane filtration methods have demonstrated recoveries ranging from approximately 10% to less than 1% during virus concentration from sewage^42^. While the BRSV recovery efficiencies in the current study span 2.7 orders of magnitude, the range of SARS-CoV-2 concentration efficiencies, only spanned 1.9 orders of magnitude.

Importantly, a statistically meaningful difference was observed in recovery efficiencies for the process control between WWTP A and B. This difference was also observed for the concentration recovery assessment of SARS-CoV-2 RNA. Enveloped viruses, such as SARS-CoV-2, partition favorably to solids^43,46,47^ and their recoveries from solids can be low^46^. The partitioning is likely further complicated by the presence of SARS-CoV-2 genetic material in various forms in wastewater, including free RNA and capsid-contained RNA^48^. In a study of SARS-CoV-2 adsorption to surfaces in solution, electrostatic adhesion correlated with both solution ionic strength and surface chemistry^49^. Additionally, a physicochemical model suggested specific absorbance of the wastewater as the parameter with the highest correlation with RNA concentration^50^. It could be expected that in wastewater, such parameters would be highly variable, and could lead to highly variable recovery efficiencies both within and between WWTPs. A negative correlation between solids and recovery efficiency was observed for electronegative membrane concentration^18^ in a WBE study in Wisconsin. This is consistent with the improved recoveries observed for BRSV after solids were removed. Despite the low and variable recovery in the current study, adsorption-extraction using electronegative membranes demonstrated reproducibility and sensitivity at reasonable cost in two previous methods comparisons^42,51^.

### SARS-CoV-2, PMMoV, and Process Control RNA Persistence

SARS-CoV-2, BRSV, and PMMoV RNA persistence was assessed for seven days under varying storage conditions to inform sample storage and handling recommendations. As shown in Figure S5A, there was a slight increase in SARS-CoV-2 RNA N1 copy number over seven days at 4°C. BRSV RNA also demonstrated a similar increase over seven days at 4°C (Figure S5C). At 25°C, N1 copy numbers increased from time zero to 24 hours and then decreased slightly over the remaining observations. A similar trend simultaneously observed for BRSV RNA suggests improved process efficiency at 24 hours followed by decreasing recovery and/or decay thereafter. PMMoV RNA copy numbers, shown in Figure S5B, displayed no appreciable decay throughout the entire 7-day experiment at both 4°C and 25°C. During pasteurization, there was no appreciable decrease of N1, PMMoV, or BRSV RNA copy numbers (Figure S6). However, over three freeze-thaw cycles, both the N1 and PMMoV RNA copy numbers decreased (Figure S7 A and B). After three freeze-thaw cycles, both SARS-CoV-2 RNA replicates were below the N1 95% LOD of 3.3 (95% CI: 2.8 – 3.8) copies per reaction (Figure S8). BRSV RNA was detectable through all three free-thaw cycles without a consistent increasing or decreasing trend (Figure S7 C).

Consistent with previous reports, SARS-CoV-2 RNA exhibited little decay in primary influent stored over seven days at both 4°C and 25°C^52–54^. However, freeze-thaw cycles degraded both SARS-CoV-2 and PMMoV RNA copy numbers. Freezing at −20°C and −80°C has been reported to decrease copy numbers for assays targeting the SARS-CoV-2 N gene^48,54^. Based on these observations, short-term storage of primary influent samples at 4°C prior to processing is preferable to freezing at −80°C. While SARS-CoV-2 RNA did not appear to decay substantially at 25°C, these observations in primary influent should not be extended to RNA persistence in raw sewage as it travels through the wastewater collection system. During pasteurization, SARS-CoV-2 RNA in primary influent persisted while infectious SARS-CoV-2 was rapidly inactivated at 50°C and 70°C^53^. Others have reported no effect of pasteurization on SARS-CoV-2 RNA copy numbers^42^ or even increases in copy number associated with pasteurization^55^. Together these observations indicate pasteurization is a reasonable biosafety strategy to mitigate infection risks associated with infectious SARS-CoV-2 while preserving genetic signal for SARS-CoV-2 RNA surveillance.

### COVID-19 Clinical Surveillance During 24-hour Sampling

Clinical COVID-19 data (both cases and percent positivity) were assessed for the two weeks prior to each sampling event. During the two weeks prior to the June 18 to 19 sampling in the county containing WWTP A, there was an average of 65.1 (95% CI: 50.5 – 79.8) daily new cases of COVID-19 and a 12.8% positivity rate (95% CI: 11.0 – 14.6). Prior to the December 2 sampling event in the same county daily new cases of COVID-19 averaged 242 (95% CI: 199 – 285) and daily positivity was 24.3% (95% CI: 20.4 – 28.2). In the county containing WWTP B from 25 April to 8 May an average of 16.4 (95% CI: 11.2 – 21.7) daily new cases and 7.0% positivity (95% CI: 5.42 – 8.51) were observed, and from 19 November to 2 December an average of 278 (95% CI: 180 – 376) daily new cases and 16.8% positivity (95% CI: 14.3 – 19.3) were recorded. The clinical COVID-19 trends in each county are illustrated in Figures S9 and S10. Statistically significant differences were inconsistent between adjacent pairs along the COVID-19 case and positivity gradient. The average daily COVID-19 cases and positivity were not statistically different between WWTP A and WWTP B during the May/June sampling (*p* = 0.1762, *p* = 0.0987, respectively), nor during the December sampling (*p* >0.9999, *p* = 0.1761, respectively). However, average COVID-19 cases were significantly less at WWTP A in June than in December (*p* = 0.0038). There was not a significant difference in the positivity rate between WWTP A and WWTP B in December (*p* = 0.4868). These data indicate that although there was a gradient in average daily COVID-19 cases and positivity between sampling events, there was not a difference in the COVID-19 clinical status within the counties containing WWTP A and WWTP B in the two weeks prior to sampling in May/June and in the two weeks prior to sampling in December. There was, however, a significant increase in COVID-19 cases, but not positivity, between May/June and December in both counties indicating that clinical surveillance systems were testing a larger number of residents by December.

### Within-Day Variation in Primary Influent

During both 24-hour sampling intervals at WWTP A (Figure S11), hourly flow rates peaked from roughly 9:00 to midnight. At WWTP B, elevated hourly flows occurred from roughly mid-day (11:00–13:00) to midnight. Flow rates were not different between sampling days for either WWTP (*p* >0.9999). Summary statistics for each parameter measured during the 24-hour sampling events are listed in Table S2. At both WWTP A and B, higher PMMoV concentrations primarily corresponded with periods of increased influent flow rate as illustrated in panels C & D of Figures S11 and S12. PMMoV concentrations in primary influent at WWTP A were comparable between sampling events (*p* >0.9999), while PMMoV concentrations were greater during the May than December sampling in primary influent at WWTP B (*p* <0.0001). For two of the four sampling days across both WWTPs, the daily time-based composites yielded lower PMMoV concentrations than the average of the grab samples. The recovery of the process control from primary influent at WWTP A was higher during periods of lower flow and PMMoV concentration (Figure S11 A & B), but there was no difference in recovery between the two sampling events at WWTP A (*p* = 0.5610). There was also no difference in BRSV recovery between sampling events at WWTP B (*p* = 0.4436) (Figure S12 A & B). However, during the May sampling event higher recoveries were observed during periods of both high and low flow. During the December sampling at WWTP B, BRSV recovery followed a pattern more similar to that observed at WWTP A with highest recoveries during periods of lower flow and PMMoV concentration.

As shown in Figure 1 A & B, SARS-CoV-2 RNA was detected in every primary influent grab sample and composite sample during both sampling events at WWTP A. The highest SARS-CoV-2 RNA concentrations were observed during overnight low-flow periods and in the morning. After recovery correction of SARS-CoV-2 RNA concentrations, periods of increased SARS-CoV-2 RNA concentrations in the primary influent of WWTP A better aligned with increased PMMoV concentrations. During the May sampling event at WWTP B, SARS-CoV-2 RNA was only detected in 6 of 12 primary influent grab samples, Figure 1 C, at concentrations below the 95% LOD. These detections all occurred from 7:00 to 21:00 with no detections overnight. During the December sampling at WWTP B (Figure 1 D), SARS-CoV-2 RNA was detected in all grab samples and the daily composite sample and the highest concentrations were observed in the morning and mid-day hours (7:00 – 15:00). Recovery adjustment of SARS-CoV-2 RNA concentrations in WWTP B primary influent accentuated the existing high concentrations during periods with high PMMoV, but did not reshape temporal trends as dramatically as the recovery adjustment at WWTP A. For all sampling days, daily time-based composite samples yielded lower SARS-CoV-2 RNA concentrations than the average of the grab samples. Process recovery efficiencies were generally lower for composite samples (0.12%, 0.17%, 0.20%, 1.25%) than for the corresponding daily average recovery among grab samples (Table S2). Even with recovery adjustment, composite samples still yielded lower SARS-CoV-2 RNA concentrations than the 24-hour average from grab samples. SARS-CoV-2 RNA trends in primary influent were also assessed using the product of the SARS-CoV-2 RNA concentration and the hourly flow rate, termed the SARS-CoV-2 RNA load in primary influent. At both WWTPs, changes in the SARS-CoV-2 RNA within-day trends mediated by flow rate were not as large as the changes mediated by recovery adjustment (Figure S13 A & B, Figure S14 A & B). The ratio of SARS-CoV-2 RNA to PMMoV RNA in log_10_ copy number per liter in primary influent at WWTP A (Figure S13 C & D) and WWTP B (Figure S14 C & D) showed similar within-day trends to the unadjusted SARS-CoV-2 RNA concentration.

**Figure 1.**
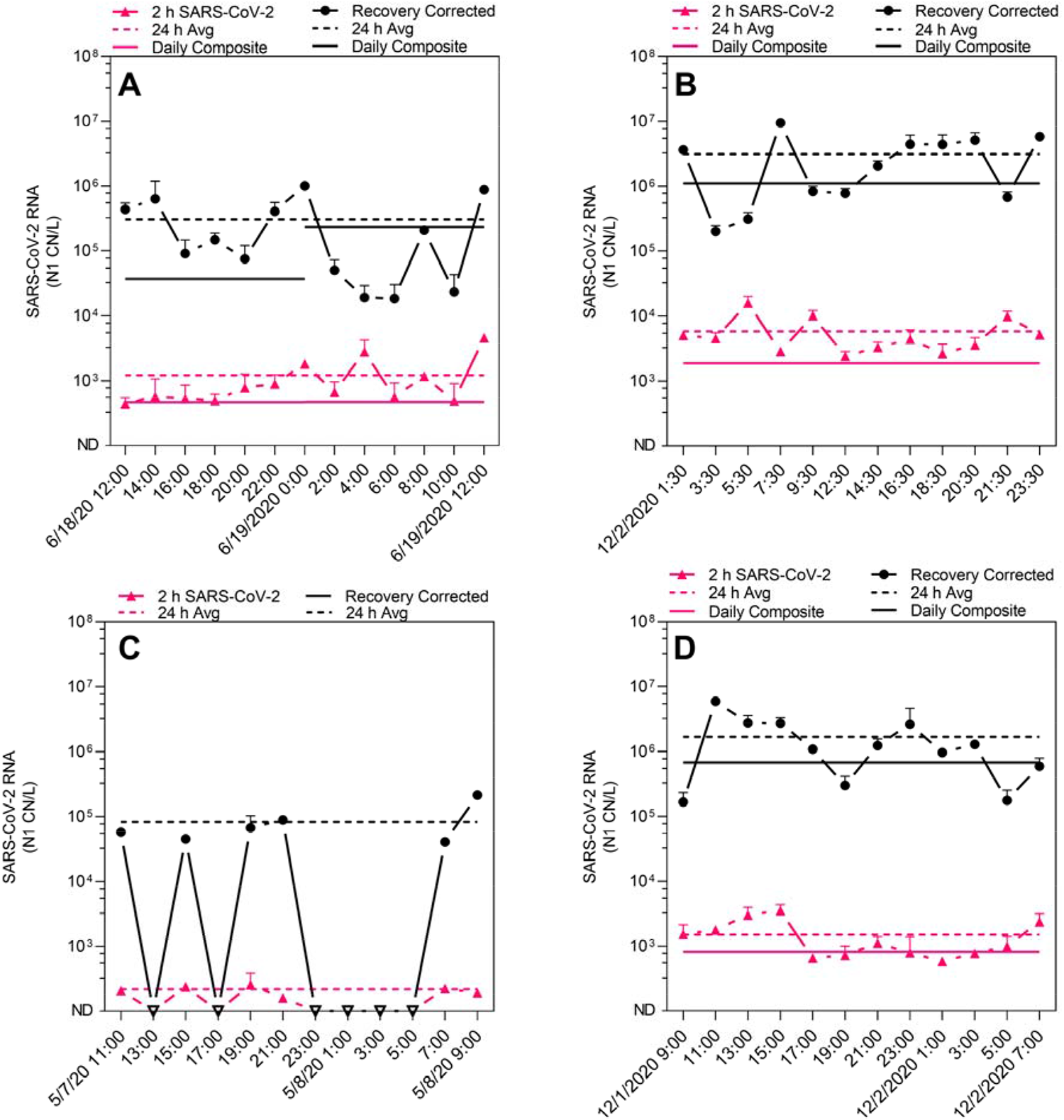
SARS-CoV-2 RNA concentrations, N1 copy number (CN) per liter, and recovery-corrected concentrations as observed in grab samples and daily composite samples of primary influent during four 24-hour sampling events at two WWTPs: June 18 to 19 at WWTP A (A), December 2 at WWTP A (B), May 7 to 8 at WWTP B (C), and December 1 to 2 at WWTP B (D).

Across all sampling periods at both WWTPs, the lowest variance was observed in the influent flow rates (CV: 7.97% - 17.1%). The greatest variance was observed for the process control recovery efficiencies (CV: 50.9% - 206%). The large variation in recovery efficiency observed during 24-hour sampling periods draws further attention to the importance the consistent use of process controls in wastewater surveillance despite their limitations^45^. Following the suggestion of Kantor *et al*., both the directly observed concentration data and recovery efficiency are reported herein along with the recovery-corrected data. The recovery adjusted SARS-CoV-2 RNA data better reflected increased SARS-CoV-2 concentration during periods of increased influent flow and fecal-indicator virus concentration. The sporadic use of process or molecular controls observed in the WBE literature greatly limits the ability to compare SARS-CoV-2 RNA measurements within and between WWTPs^56^. The observations in this study reinforce that consistent assessment of process recovery efficiency via appropriate controls is a vital component of wastewater surveillance for SARS-CoV-2 RNA.

In primary influent PMMoV RNA concentrations exhibited greater mean concentration (6.6 – 7.1 log_10_ CN/L) and lower variance (CV: 34.9 – 67.5%) than SARS-CoV-2 RNA concentrations (mean: 3.1 log_10_ – 3.8 log_10_ CN/L; CV: 70 – 100%). Increased mean concentration and lower variance in primary influent, consistent with other observations of primary influent^57,58^, is expected given the likely greater prevalence of PMMoV RNA shedding^23^ compared to SARS-CoV-2 RNA shedding among the sewershed population. Similar trends between PMMoV and human adenovirus DNA were observed during 24-hour sampling of primary influent from WWTPs in Australia^59^. Unlike the studies in Nevada^28^ and Virginia^60^, PMMoV RNA and SARS-CoV-2 RNA concentrations in 24-hour time-based composite samples were frequently lower than the grab sample derived average. The process control recoveries were also generally lower for the composite samples. Interpreted together, the low SARS-CoV-2 RNA concentrations using time-based composites in the current study combined with the similar and higher concentrations using flow-weighted composites in the previous studies strongly suggest diurnal variation in SARS-CoV-2 RNA concentrations in primary influent. Recovery adjustment increased the variation in SARS-CoV-2 concentrations more greatly than flow adjustment (CV: 16.3 – 104% compared to 11 – 76%, respectively). Normalization of SARS-CoV-2 RNA concentrations using PMMoV, as has been done for both primary influent^18^ and primary solids^8,61^, greatly reduced the observed variation (CV: 4 – 13%). For comparing to clinical surveillance data, variations in recovery efficiency likely represent a significant covariant^62^, but normalization by PMMoV may greatly reduce the variability of the genetic signal in wastewater.

### WWTP Influent SARS-CoV-2 RNA and Clinical Surveillance

Due to the agreement between BRSV and SARS-CoV-2 RNA recovery, the effect of recovery-correction on within-day trends, and the large variance associated with recovery efficiency, recovery-corrected SARS-CoV-2 RNA concentrations in primary influent were compared to county-level COVID-19 cases and positivity rates during the two weeks prior to each 24-hour sampling period. SARS-CoV-2 RNA concentrations (Figure 2A) did not consistently increase with increasing average daily COVID-19 cases. The observed non-linear trend was also observed between PMMoV-normalized SARS-CoV-2 RNA concentrations (Figure 2C). For average daily COVID-19 positivity (Figure 2B) there was ordinal agreement between positivity and mean SARS-CoV-2 RNA concentrations in primary influent. However, after accounting for within-day variation in SARS-CoV-2 concentrations and recovery, the statistical differences between SARS-CoV-2 RNA concentrations and COVID-19 positivity were only consistent for two of three increases. As shown in Figure 2D, a similar trend was observed between the ordinal agreement of PMMoV-normalized SARS-CoV-2 RNA concentrations and COVID-19 positivity. Trends between unadjusted SARS-CoV-2 RNA concentrations and clinical surveillance data, which do not demonstrate improved agreement, are shown in Figure S15.

**Figure 2.**
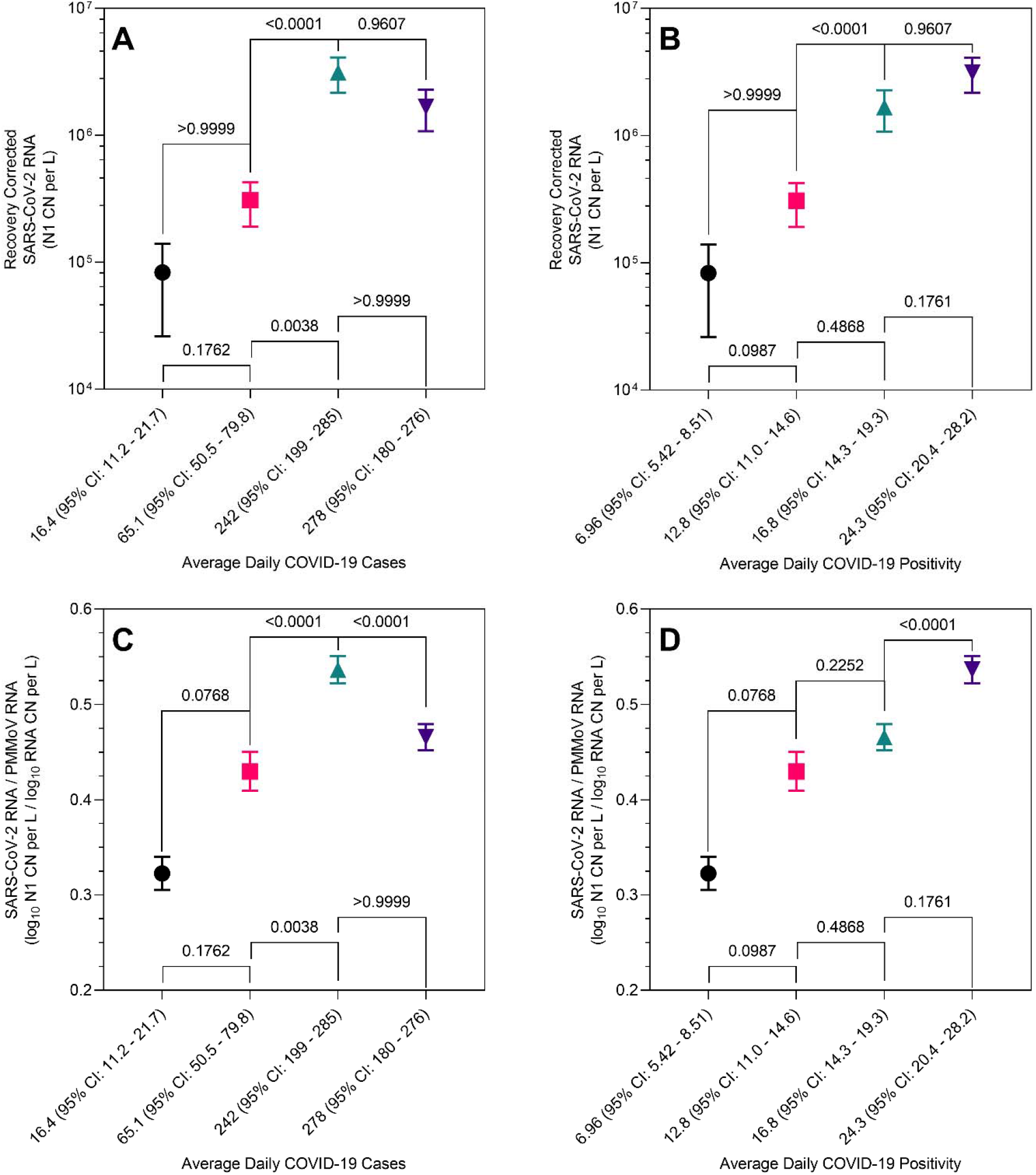
Recovery corrected SARS-CoV-2 RNA concentrations, N1 copy number (CN) per liter, in primary influent stratified by increasing average daily COVID-19 cases (A) and average daily COVID-19 positivity (B) and PMMoV-normalized SARS-CoV-2 RNA concentrations (log_10_N1 CN per L/log_10_ CN per L) in primary influent by increasing average daily COVID-19 cases (C) and average daily COVID-19 positivity (D). COVID-19 case and positivity averages and confidence intervals are calculated for the two-week period prior to each 24-h sampling event. Statistical comparisons between adjacent pairs were made using Kruskal-Wallis tests with Dunn’s correction.

Both recovery-corrected SARS-CoV-2 RNA concentrations and PMMoV-normalized SARS-CoV-2 RNA concentrations in primary influent demonstrated non-linear trends with county-level average daily COVID-19 cases. However, county-level COVID-19 positivity showed ordinal agreement with both SARS-CoV-2 RNA concentration (both corrected and uncorrected for recovery) and PMMoV-normalized SARS-CoV-2 RNA concentration in primary influent. Given that each of the WWTPs in the current study are located in counties with large peri-urban and rural areas and large portions of the population living outside the sewershed or connected to on-site septic systems, it is reasonable that positivity in COVID-19 testing better reflects primary influent concentrations than total new COVID-19 cases for the county. Positivity at the county-level is more likely to represent the clinical trends within the sewershed since it accounts for the number of cases, the number of tests administered, and presence of unidentified infections. Correlations have been demonstrated between COVID-19 cases within sewershed boundaries and SARS-CoV-2 RNA in wastewater^17,18^, primary solids^7,8^, and between PMMoV-normalized SARS-CoV-2 RNA in primary solids^26^. SARS-CoV-2 RNA concentrations in wastewater have also been found to correlate with positivity^19^. The results of the current study indicate that when sub-county level COVID-19 clinical surveillance data are not available, positivity may offer a better metric for comparison with wastewater data.

Despite the ordinal agreement between positivity and the average concentrations in the primary influent, differences in the primary influent concentrations during each of these periods were often not meaningfully different after accounting for variation in concentration and recovery. A Bayesian modeling experiment indicated that variation in recovery efficiency is a key constraint in using wastewater data to estimate prevalence^62^. These observations indicate that quantitative relationships between wastewater data and SARS-CoV-2 infection prevalence, particularly those premised on material balance, are likely to remain constrained by variability and uncertainty^21^.

There are several limitations for the current study. The work included only two WWTPs in northern Indiana, USA sampled over two 24-hour periods each. These WWTPs are located in counties that include large rural and peri-urban areas with many residents connected to septic systems and may not be generalized to all sewersheds particularly urban ones. The four 24-hour sampling periods spanned weekday periods from Tuesday to Wednesday and Thursday to Friday and do not include weekend periods. The sewershed served by WWTP A, where large variations in process recovery were observed, includes large manufacturing and industrial areas characterized by 24-hour shift work. Additionally, flow patterns during all the sampling events were likely affected by changes in human behavior patterns associated with lockdowns and interrupted domestic and working routines. The concentration method utilized to detect and quantify RNA present at low levels was characterized by low and variable recovery in the course of the study. This injects additional variation and uncertainty into the trends that were observed. Even so, recovery-corrected SARS-CoV-2 RNA concentrations reflected within-day trends in influent flow rate and fecal-indicator virus concentrations. Both recovery-corrected and PMMoV-normalized SARS-CoV-2 concentrations in primary influent demonstrated increases that were consistent with ordinal increases in COVID-19 positivity prior to each of the 24-hour sampling periods. These findings indicate the genetic signal in primary influent from two WWTPs, both in rural and peri-urban counties, reflects increasing COVID-19 positivity. In such communities, where clinical surveillance capacity might be limited, WBE shows potential for monitoring SARS-CoV-2 infection and COVID-19 trends.

## Data Availability

Data available in SI.

## AUTHOR’S STATEMENT

The authors declare no competing financial interest.

## ACKNOWLEDGMENTS

This study was funded by United States National Science Foundation under grant 2027752. This study would not be possible without the gracious support of our municipal utility partners who collected the primary influent samples used in this study and provided access to their facilities.

## REFERENCES

(1) Lee, S.; Kim, T.; Lee, E.; Lee, C.; Kim, H.; Rhee, H.; Park, S. Y.; Son, H.-J.; Yu, S.; Park, J. W.; … Kim, T. H. Clinical Course and Molecular Viral Shedding Among Asymptomatic and Symptomatic Patients With SARS-CoV-2 Infection in a Community Treatment Center in the Republic of Korea. JAMA Intern. Med. 2020, 180 (11), 1447. https://doi.org/10.1001/jamainternmed.2020.3862.

(2) Wölfel, R.; Corman, V. M.; Guggemos, W.; Seilmaier, M.; Müller, M. A.; Niemeyer, D.; Kelly, T. C. J.; Vollmar, P.; Hoelscher, M.; Bleicker, T.; … Bleicker, T. Virological Assessment of Hospitalized Cases of Coronavirus Disease 2019. medRxiv 2020.

(3) He, X.; Lau, E. H. Y.; Wu, P.; Deng, X.; Wang, J.; Hao, X.; Lau, Y. C.; Wong, J. Y.; Guan, Y.; Tan, X.; … Leung, G. M. Temporal Dynamics in Viral Shedding and Transmissibility of COVID-19. Nat. Med. 2020, 26 (5), 672–675. https://doi.org/10.1038/s41591-020-0869-5.

(4) Li, W.; Su, Y.-Y.; Zhi, S.-S.; Huang, J.; Zhuang, C.-L.; Bai, W.-Z.; Wan, Y.; Meng, X.-R.; Zhang, L.; Zhou, Y.-B.; … Ma, Y. Virus Shedding Dynamics in Asymptomatic and Mildly Symptomatic Patients Infected with SARS-CoV-2. Clin. Microbiol. Infect. 2020, 26 (11), 1556.e1-1556.e6. https://doi.org/10.1016/j.cmi.2020.07.008.

(5) Bivins, A.; North, D.; Ahmad, A.; Ahmed, W.; Alm, E.; Been, F.; Bhattacharya, P.; Bijlsma, L.; Boehm, A. B.; Brown, J.; … Bibby, K. Wastewater-Based Epidemiology: Global Collaborative to Maximize Contributions in the Fight against COVID-19. Environ. Sci. Technol. 2020, No. Figure 1. https://doi.org/10.1021/acs.est.0c02388.

(6) Medema, G.; Heijnen, L.; Elsinga, G.; Italiaander, R.; Brouwer, A. Presence of SARS-Coronavirus-2 RNA in Sewage and Correlation with Reported COVID-19 Prevalence in the Early Stage of the Epidemic in the Netherlands. 2020. https://doi.org/10.1021/acs.estlett.0c00357.

(7) Peccia, J.; Zulli, A.; Brackney, D. E.; Grubaugh, N. D.; Edward, H.; Casanovas-massana, A.; Ko, A. I.; Malik, A. A.; Wang, D. SARS-CoV-2 RNA Concentrations in Primary Municipal Sewage Sludge as a Leading Indicator of COVID-19 Outbreak Dynamics. 2020, 1 (203).

(8) Graham, K. E.; Loeb, S. K.; Wolfe, M. K.; Catoe, D.; Sinnott-Armstrong, N.; Kim, S.; Yamahara, K. M.; Sassoubre, L. M.; Mendoza Grijalva, L. M., Roldan-Hernandez, L.; … Boehm, A. B. SARS-CoV-2 RNA in Wastewater Settled Solids Is Associated with COVID-19 Cases in a Large Urban Sewershed. Environ. Sci. Technol. 2021, 55 (1), 488–498. https://doi.org/10.1021/acs.est.0c06191.

(9) Gonzalez, R.; Curtis, K.; Bivins, A.; Bibby, K.; Weir, M. H.; Yetka, K.; Thompson, H.; Keeling, D.; Mitchell, J.; Gonzalez, D. COVID-19 Surveillance in Southeastern Virginia Using Wastewater-Based Epidemiology. Water Res. 2020, 186, 116296. https://doi.org/10.1016/j.watres.2020.116296.

(10) Hemalatha, M.; Kiran, U.; Kuncha, S. K.; Kopperi, H.; Gokulan, C. G.; Mohan, S. V.; Mishra, R. K. Surveillance of SARS-CoV-2 Spread Using Wastewater-Based Epidemiology: Comprehensive Study. Sci. Total Environ. 2021, 768, 144704. https://doi.org/10.1016/j.scitotenv.2020.144704.

(11) Hata, A.; Hara-Yamamura, H.; Meuchi, Y.; Imai, S.; Honda, R. Detection of SARS-CoV-2 in Wastewater in Japan during a COVID-19 Outbreak. Sci. Total Environ. 2021, 758, 143578. https://doi.org/10.1016/j.scitotenv.2020.143578.

(12) Randazzo, W.; Truchado, P.; Cuevas-Ferrando, E.; Simón, P.; Allende, A.; Sánchez, G. SARS-CoV-2 RNA in Wastewater Anticipated COVID-19 Occurrence in a Low Prevalence Area. Water Res. 2020, 181, 115942. https://doi.org/10.1016/j.watres.2020.115942.

(13) Emerging Issues in the Water Environment during Anthropocene: A South Asian Perspective; Kumar, M.; Snow, D. D.; Honda, R.; Eds., Springer: Singapore, 2020. https://doi.org/10.1007/978-981-32-9771-5.

(14) Kumar, M.; Patel, A. K.; Shah, A. V.; Raval, J.; Rajpara, N.; Joshi, M.; Joshi, C. G. First Proof of the Capability of Wastewater Surveillance for COVID-19 in India through Detection of Genetic Material of SARS-CoV-2. Sci. Total Environ. 2020, 746, 141326. https://doi.org/10.1016/j.scitotenv.2020.141326.

(15) La Rosa, G.; Iaconelli, M.; Mancini, P.; Bonanno Ferraro, G.; Veneri, C.; Bonadonna, L.; Lucentini, L.; Suffredini, E. First Detection of SARS-CoV-2 in Untreated Wastewaters in Italy. Sci. Total Environ. 2020, 736, 139652. https://doi.org/10.1016/j.scitotenv.2020.139652.

(16) Nemudryi, A.; Nemudraia, A.; Wiegand, T.; Surya, K.; Buyukyoruk, M.; Cicha, C.; Vanderwood, K. K.; Wilkinson, R.; Wiedenheft, B. Temporal Detection and Phylogenetic Assessment of SARS-CoV-2 in Municipal Wastewater. Cell Reports Med. 2020, 1 (6), 100098. https://doi.org/10.1016/j.xcrm.2020.100098.

(17) Weidhaas, J.; Aanderud, Z. T.; Roper, D. K.; VanDerslice, J.; Gaddis, E. B.; Ostermiller, J.; Hoffman, K.; Jamal, R.; Heck, P.; Zhang, Y.; … LaCross, N. Correlation of SARS-CoV-2 RNA in Wastewater with COVID-19 Disease Burden in Sewersheds. Sci. Total Environ. 2021, 775, 145790. https://doi.org/10.1016/j.scitotenv.2021.145790.

(18) Feng, S.; Roguet, A.; McClary-Gutierrez, J. S.; Newton, R. J.; Kloczko, N.; Meiman, J. G.; McLellan, S. L. Evaluation of Sampling Frequency and Normalization of SARS-CoV-2 Wastewater Concentrations for Capturing COVID-19 Burdens in the Community. medRxiv 2021, preprint. https://doi.org/10.1101/2021.02.17.21251867v2.

(19) Stadler, L. B.; Ensor, K. B.; Clark, J. R.; Kalvapalle, P.; LaTurner, Z. W.; Mojica, L.; Terwilliger, A.; Zhuo, Y.; Ali, P.; Avadhanula, V.; … Hopkins, L. Wastewater Analysis of SARS-CoV-2 as a Predictive Metric of Positivity Rate for a Major Metropolis. medRxiv 2020, preprint. https://doi.org/10.1101/2020.11.04.20226191v1.

(20) Wu, F.; Zhang, J.; Xiao, A.; Gu, X.; Lee, W. L.; Armas, F.; Kauffman, K.; Hanage, W.; Matus, M.; Ghaeli, N.; … Alm, E. J. SARS-CoV-2 Titers in Wastewater Are Higher than Expected from Clinically Confirmed Cases. mSystems 2020, 5 (4). https://doi.org/10.1128/mSystems.00614-20.

(21) Ahmed, W.; Angel, N.; Edson, J.; Bibby, K.; Bivins, A.; O’Brien, J. W.; Choi, P. M.; Kitajima, M.; Simpson, S. L.; Li, J.; … Mueller, J. F. First Confirmed Detection of SARS-CoV-2 in Untreated Wastewater in Australia: A Proof of Concept for the Wastewater Surveillance of COVID-19 in the Community. Sci. Total Environ. 2020, 138764. https://doi.org/10.1016/j.scitotenv.2020.138764.

(22) Bivins, A.; Crank, K.; Greaves, J.; North, D.; Wu, Z.; Bibby, K. CrAssphage and Pepper Mild Mottle Virus as Viral Water Quality Monitoring Tools – Potential, Research Gaps, and Way Forward. Curr. Opin. Environ. Sci. Heal. 2020. https://doi.org/https://doi.org/10.1016/j.coesh.2020.02.001.

(23) Zhang, T.; Breitbart, M.; Lee, W. H.; Run, J. Q.; Wei, C. L.; Soh, S. W. L.; Hibberd, M. L.; Liu, E. T.; Rohwer, F.; Ruan, Y. RNA Viral Community in Human Feces: Prevalence of Plant Pathogenic Viruses. PLoS Biol. 2006, 4 (1), 0108–0118. https://doi.org/10.1371/journal.pbio.0040003.

(24) Rosario, K.; Symonds, E. M.; Sinigalliano, C.; Stewart, J.; Breitbart, M. Pepper Mild Mottle Virus as an Indicator of Fecal Pollution. Appl. Environ. Microbiol. 2009, 75 (22), 7261–7267. https://doi.org/10.1128/AEM.00410-09.

(25) Symonds, E. M.; Rosario, K.; Breitbart, M. Pepper Mild Mottle Virus: Agricultural Menace Turned Effective Tool for Microbial Water Quality Monitoring and Assessing (Waste)Water Treatment Technologies. PLoS Pathog. 2019, 15 (4), e1007639. https://doi.org/10.1371/journal.ppat.1007639.

(26) D’Aoust, P. M.; Mercier, E.; Montpetit, D.; Jia, J.-J.; Alexandrov, I.; Neault, N.; Baig, A. T.; Mayne, J.; Zhang, X.; Alain, T.; … Delatolla, R. Quantitative Analysis of SARS-CoV-2 RNA from Wastewater Solids in Communities with Low COVID-19 Incidence and Prevalence. Water Res. 2021, 188, 116560. https://doi.org/10.1016/j.watres.2020.116560.

(27) Curtis, K.; Keeling, D.; Yetka, K.; Larson, A.; Gonzalez, R. Wastewater SARS-CoV-2 Concentration and Loading Variability from Grab and 24-Hour Composite Samples. medRxiv 2020, preprint. https://doi.org/10.1101/2020.07.10.20150607v1.

(28) Gerrity, D.; Papp, K.; Stoker, M.; Sims, A.; Frehner, W. Early-Pandemic Wastewater Surveillance of SARS-CoV-2 in Southern Nevada: Methodology, Occurrence, and Incidence/Prevalence Considerations. Water Res. X 2021, 10, 100086. https://doi.org/10.1016/j.wroa.2020.100086.

(29) Ahmed, W.; Harwood, V. J.; Gyawali, P.; Sidhu, J. P. S.; Toze, S. Comparison of Concentration Methods for Quantitative Detection of Sewage-Associated Viral Markers in Environmental Waters. Appl. Environ. Microbiol. 2015, 81 (6), 2042–2049. https://doi.org/10.1128/AEM.03851-14.

(30) Valarcher, J.-F.; Taylor, G. Bovine Respiratory Syncytial Virus Infection. Vet. Res. 2007, 38 (2), 153–180. https://doi.org/10.1051/vetres:2006053.

(31) Cashdollar, J. L.; Brinkman, N. E.; Griffin, S. M.; McMinn, B. R.; Rhodes, E. R.; Varughese, E. A.; Grimm, A. C.; Parshionikar, S. U.; Wymer, L.; Fout, G. S. Development and Evaluation of EPA Method 1615 for Detection of Enterovirus and Norovirus in Water. Appl. Environ. Microbiol. 2013, 79 (1), 215–223. https://doi.org/10.1128/AEM.02270-12.

(32) Lu, X.; Wang, L.; Sakthivel, S. K.; Whitaker, B.; Murray, J.; Kamili, S.; Lynch, B.; Malapati, L.; Burke, S. A.; Harcourt, J.; … Lindstrom, S. US CDC Real-Time Reverse Transcription PCR Panel for Detection of Severe Acute Respiratory Syndrome Coronavirus 2. Emerg. Infect. Dis. 2020, 26 (8), 1654–1665. https://doi.org/10.3201/eid2608.201246.

(33) Haramoto, E.; Kitajima, M.; Kishida, N.; Konno, Y.; Katayama, H.; Asami, M.; Akiba, M. Occurrence of Pepper Mild Mottle Virus in Drinking Water Sources in Japan. Appl. Environ. Microbiol. 2013, 79 (23), 7413–7418. https://doi.org/10.1128/AEM.02354-13.

(34) Boxus, M.; Letellier, C.; Kerkhofs, P. Real Time RT-PCR for the Detection and Quantitation of Bovine Respiratory Syncytial Virus. J. Virol. Methods 2005, 125 (2), 125–130. https://doi.org/10.1016/j.jviromet.2005.01.008.

(35) Bivins, A.; Lowry, S.; Murphy, H. M.; Borchardt, M.; Coyte, R.; Labhasetwar, P.; Brown, J. Waterborne Pathogen Monitoring in Jaipur, India Reveals Potential Microbial Risks of Urban Groundwater Supply. npj Clean Water 2020, 3 (1), 35. https://doi.org/10.1038/s41545-020-00081-3.

(36) Dunn, O. J. Multiple Comparisons Using Rank Sums. Technometrics 1964, 6 (3), 241–252. https://doi.org/10.1080/00401706.1964.10490181.

(37) Kruskal, W. H.; Wallis, W. A. Use of Ranks in One-Criterion Variance Analysis. J. Am. Stat. Assoc. 1952, 47 (260), 583–621. https://doi.org/10.1080/01621459.1952.10483441.

(38) Mann, H. B.; Whitney, D. R. On a Test of Whether One of Two Random Variables Is Stochastically Larger than the Other. Ann. Math. Stat. 1947, 18 (1), 50–60. https://doi.org/10.1214/aoms/1177730491.

(39) Sherchan, S. P.; Shahin, S.; Ward, L. M.; Tandukar, S.; Aw, T. G.; Schmitz, B.; Ahmed, W.; Kitajima, M. First Detection of SARS-CoV-2 RNA in Wastewater in North America: A Study in Louisiana, USA. Sci. Total Environ. 2020, 743, 140621. https://doi.org/10.1016/j.scitotenv.2020.140621.

(40) Ahmed, W.; Bertsch, P. M.; Bivins, A.; Bibby, K.; Farkas, K.; Gathercole, A.; Haramoto, E.; Gyawali, P.; Korajkic, A.; McMinn, B. R.; … Kitajima, M. Comparison of Virus Concentration Methods for the RT-QPCR-Based Recovery of Murine Hepatitis Virus, a Surrogate for SARS-CoV-2 from Untreated Wastewater. Sci. Total Environ. 2020, 739 (June), 139960. https://doi.org/10.1016/j.scitotenv.2020.139960.

(41) Farkas, K.; Hillary, L. S.; Thorpe, J.; Walker, D. I.; Lowther, J. A.; McDonald, J. E.; Malham, S. K.; Jones, D. L. Concentration and Quantification of SARS-CoV-2 RNA in Wastewater Using Polyethylene Glycol-Based Concentration and QRT-PCR. Methods Protoc. 2021, 4 (1), 17. https://doi.org/10.3390/mps4010017.

(42) Pecson, B. M.; Darby, E.; Haas, C. N.; Amha, Y. M.; Bartolo, M.; Danielson, R.; Dearborn, Y.; Di Giovanni, G.; Ferguson, C.; Fevig, S.; … SARS-CoV-2 Interlaboratory Consortium. Reproducibility and Sensitivity of 36 Methods to Quantify the SARS-CoV-2 Genetic Signal in Raw Wastewater: Findings from an Interlaboratory Methods Evaluation in the U.S. Environ. Sci. Water Res. Technol. 2021. https://doi.org/10.1039/D0EW00946F.

(43) Chik, A. H. S.; Glier, M. B.; Servos, M.; Mangat, C. S.; Pang, X.-L.; Qiu, Y.; D’Aoust, P. M.; Burnet, J.-B.; Delatolla, R.; Dorner, S.; … Hrudey, S. E. Comparison of Approaches to Quantify SARS-CoV-2 in Wastewater Using RT-QPCR: Results and Implications from a Collaborative Inter-Laboratory Study in Canada. J. Environ. Sci. 2021, 107, 218–229. https://doi.org/10.1016/j.jes.2021.01.029.

(44) Karthikeyan, S.; Ronquillo, N.; Belda-Ferre, P.; Alvarado, D.; Javidi, T.; Longhurst, C. A.; Knight, R. High-Throughput Wastewater SARS-CoV-2 Detection Enables Forecasting of Community Infection Dynamics in San Diego County. mSystems 2021, 6 (2). https://doi.org/10.1128/mSystems.00045-21.

(45) Kantor, R. S.; Nelson, K. L.; Greenwald, H. D.; Kennedy, L. C. Challenges in Measuring the Recovery of SARS-CoV-2 from Wastewater. Environ. Sci. Technol. 2021, acs.est.0c08210. https://doi.org/10.1021/acs.est.0c08210.

(46) Ye, Y.; Ellenberg, R. M.; Graham, K. E.; Wigginton, K. R. Survivability, Partitioning, and Recovery of Enveloped Viruses in Untreated Municipal Wastewater. Environ. Sci. Technol. 2016, 50 (10), 5077–5085. https://doi.org/10.1021/acs.est.6b00876.

(47) Balboa, S.; Mauricio-Iglesias, M.; Rodriguez, S.; Martínez-Lamas, L.; Vasallo, F. J.; Regueiro, B.; Lema, J. M. The Fate of SARS-COV-2 in WWTPS Points out the Sludge Line as a Suitable Spot for Detection of COVID-19. Sci. Total Environ. 2021, 772, 145268. https://doi.org/10.1016/j.scitotenv.2021.145268.

(48) Wurtzer, S.; Waldman, P.; Ferrier-Rembert, A.; Frenois-Veyrat, G.; Mouchel, J. M.; Boni, M.; Maday, Y.; OBEPINE Consortium; Marechal; V., Moulin, L. Several Forms of SARS-CoV-2 RNA Can Be Detected in Wastewatersl: Implication for Wastewater-Based Epidemiology and Risk Assessment. medRxiv 2020, preprint. https://doi.org/10.1101/2020.12.19.20248508v1.

(49) Liu, Y.-N.; Lv, Z.-T.; Yang, S.-Y.; Liu, X.-W. Optical Tracking of the Interfacial Dynamics of Single SARS-CoV-2 Pseudoviruses. Environ. Sci. Technol. 2021, acs.est.0c06962. https://doi.org/10.1021/acs.est.0c06962.

(50) Petala, M.; Dafou, D.; Kostoglou, M.; Karapantsios, T.; Kanata, E.; Chatziefstathiou, A.; Sakaveli, F.; Kotoulas, K.; Arsenakis, M.; Roilides, E.; … Papaioannou, N. A Physicochemical Model for Rationalizing SARS-CoV-2 Concentration in Sewage. Case Study: The City of Thessaloniki in Greece. Sci. Total Environ. 2021, 755, 142855. https://doi.org/10.1016/j.scitotenv.2020.142855.

(51) Zachary W. LaTurner; David M. Zong; Prashant Kalvapalle; Kiara Reyes Gamas; Austen Terwilliger; Tessa Crosby; Priyanka Ali; Vasanthi Avadhanula; Haroldo Hernandez Santos; Kyle Weesner; Loren Hopkins; Pedro A. Piedra; Anthony W. Maresso; L. B. S. Evaluating Recovery; Cost, and Throughput of Different Concentration Methods for SARS-CoV-2 Wastewater-Based Epidemiology. medRxiv 2020, preprint.

(52) Ahmed, W.; Bertsch, P. M.; Bibby, K.; Haramoto, E.; Hewitt, J.; Huygens, F.; Gyawali, P.; Korajkic, A.; Riddell, S.; Sherchan, S. P.; … Bivins, A. Decay of SARS-CoV-2 and Surrogate Murine Hepatitis Virus RNA in Untreated Wastewater to Inform Application in Wastewater-Based Epidemiology. Environ. Res. 2020, 191, 110092. https://doi.org/10.1016/j.envres.2020.110092.

(53) Bivins, A.; Greaves, J.; Fischer, R.; Yinda, K. C.; Ahmed, W.; Kitajima, M.; Munster, V. J.; Bibby, K. Persistence of SARS-CoV-2 in Water and Wastewater. Environ. Sci. Technol. Lett. 2020, acs.estlett.0c00730. https://doi.org/10.1021/acs.estlett.0c00730.

(54) Markt, R.; Mayr, M.; Peer, E.; Wagner, Andreas O.; N. L., Insam, H. Detection and Stability of SARS-CoV-2 Fragments in Wastewater: Impact of Storage Temperature. medRxiv 2021, preprint.

(55) Monica Trujillo; Kristen Cheung; Anna Gao; Irene Hoxie; Sherin Kannoly; Nanami Kubota; Kaung Myat San; D. S. S., Dennehy, J. J. Protocol for Safe, Affordable, and Reproducible Isolation and Quantitation of SARS-CoV-2 RNA from Wastewater. medRxiv 2021, preprint. https://doi.org/10.1101/2021.02.16.21251787v1.

(56) Ahmed, W.; Bivins, A.; Bertsch, P. M.; Bibby, K.; Choi, P. M.; Farkas, K.; Gyawali, P.; Hamilton, K. A.; Haramoto, E.; Kitajima, M.; … Mueller, J. F. Surveillance of SARS-CoV-2 RNA in Wastewater: Methods Optimisation and Quality Control Are Crucial for Generating Reliable Public Health Information. Curr. Opin. Environ. Sci. Heal. 2020. https://doi.org/10.1016/j.coesh.2020.09.003.

(57) Tandukar, S.; Sherchan, S. P.; Haramoto, E. Applicability of CrAssphage, Pepper Mild Mottle Virus, and Tobacco Mosaic Virus as Indicators of Reduction of Enteric Viruses during Wastewater Treatment. Sci. Rep. 2020, 10 (1), 1–8. https://doi.org/10.1038/s41598-020-60547-9.

(58) Kitajima, M.; Iker, B. C.; Pepper, I. L.; Gerba, C. P. Relative Abundance and Treatment Reduction of Viruses during Wastewater Treatment Processes - Identification of Potential Viral Indicators. Sci. Total Environ. 2014, 488–489 (1), 290–296. https://doi.org/10.1016/j.scitotenv.2014.04.087.

(59) Ahmed, W.; Bivins, A.; Bertsch, P. M.; Bibby, K.; Gyawali, P.; Sherchan, S. P.; Simpson, S. L.; Thomas, K. V.; Verhagen, R.; Kitajima, M.; … Korajkic, A. Intraday Variability of Indicator and Pathogenic Viruses in 1-h and 24-h Composite Wastewater Samples: Implications for Wastewater-Based Epidemiology. Environ. Res. 2020, 110531. https://doi.org/10.1016/j.envres.2020.110531.

(60) Curtis, K.; Keeling, D.; Yetka, K.; Larson, A.; Gonzalez, R. Wastewater SARS-CoV-2 Concentration and Loading Variability from Grab and 24-Hour Composite Samples. medRxiv 2020, 2020.07.10.20150607. https://doi.org/10.1101/2020.07.10.20150607.

(61) D’Aoust, P. M.; Graber, T. E.; Mercier, E.; Montpetit, D.; Alexandrov, I.; Neault, N.; Baig, A. T.; Mayne, J.; Zhang, X.; Alain, T.; … Delatolla, R. Catching a Resurgence: Increase in SARS-CoV-2 Viral RNA Identified in Wastewater 48 h before COVID-19 Clinical Tests and 96 h before Hospitalizations. Sci. Total Environ. 2021, 770, 145319. https://doi.org/10.1016/j.scitotenv.2021.145319.

(62) Li, X.; Zhang, S.; Shi, J.; Luby, S. P.; Jiang, G. Uncertainties in Estimating SARS-CoV-2 Prevalence by Wastewater-Based Epidemiology. Chem. Eng. J. 2021, 415, 129039. https://doi.org/10.1016/j.cej.2021.129039.

