## Supplemental Tables and Figures for "Within-Day Variability of SARS-CoV-2 RNA in Municipal Wastewater Influent During Periods of Varying COVID-19 Prevalence and Positivity"

17 pages, 2 tables, 15 figures

Table S1: RT-ddPCR assays
Figure S1: Process control recovery efficiency
Figure S2: Extraction and molecular control recovery efficiency
Figure S3: SARS-CoV-2 RNA concentration recovery efficiency
Figure S4: PMMoV RNA concentration recovery efficiency
Figure S5: RNA copy number persistence in primary influent at 4°C and 25°C
Figure S6: RNA copy number measurements before and after pasteurization
Figure S7: RNA copy number measurements following freeze-thaw cycles
Figure S8: RT-ddPCR N1 assay 95% limit of detection
Figure S9: COVID-19 clinical data from county containing WWTP A
Figure S10: COVID-19 clinical data from country containing WWTP B
Figure S11: WWTP A primary influent flow, PMMoV concentration, and recovery efficiency
Figure S12: WWTP B primary influent flow, PMMoV concentration, and recovery efficiency
Table S2: Summary statistics for primary influent flow parameters
Figure S13: WWTP A primary influent SARS-CoV-2 RNA load and PMMoV-normalized
Figure S14: WWTP B primary influent SARS-CoV-2 RNA load and PMMoV RNA-normalized
Figure S15: Primary influent SARS-CoV-2 RNA N1 concentration versus COVID-19 clinical data

Table S1 | RT-ddPCR assays used to detect and quantify SARS-CoV-2 RNA, PMMoV RNA, and BRSV RNA in primary influent wastewater samples. The assay used to quantify the extraction and molecular control Hep G armored RNA is also summarized.

| Virus | Gene Target | | Sequences | RT-ddPCR Reaction Concentration | Thermal Cycling Conditions |
|---|---|---|---|---|---|
| SARS-CoV-2 | N1 | F | 5’-GAC CCC AAA ATC AGC GAA AT-3’ | 1000 nM | 50°C 60 min; 95°C 10 min; 40 Cycles: 95°C 30 s, 59°C 60 s 98°C 10 min; 4°C hold |
| | | R | 5’-TCT GGT TAC TGC CAG TTG AAT CTG-3’ | 1000 nM | |
| | | P | 5’-FAM-ACC CCG CAT TAC GTT TGG TGG ACC-BHQ1-3’ | 250 nM | |
| PMMoV (fecal indicator virus) | replicase protein | F | 5'-GAG TGG TTT GAC CTT AAC GTT TGA-3' | 900 nM | |
| | | R | 5'-TTG TCG GTT GCA ATG CAA GT-3' | 900 nM | |
| | | P | 5'-FAM-CCT ACC GAA GCA AAT G-MGBNFQ-3' | 250 nM | |
| BRSV (process control) | nucleoprotein | F | 5'-GCA ATG CTG CAG GAC TAG GTA TAA T-3' | 900 nM | |
| | | R | 5'-ACA CTG TAA TTG ATG ACC CCA TTC T-3' | 900 nM | |
| | | P | 5’-HEX-AC CAA GAC T/ZEN/T GTA TGA TGC TGC CAA AGC A-IABkFQ-3’ | 250 nM | |
| Hep G Armored RNA (extraction & molecular control) | polyprotein precursor | F | 5’-CGG CCA AAA GGT GGT GGA TG-3’ | 900 nM | 50°C 60 min; 95°C 10 min; 40 Cycles: 95°C 30 s, 55°C 60 s 98°C 10 min; 4°C hold |
| | | R | 5’-CCC GAC GTC AGG CTC GTC G-3’ | 900 nM | |
| | | P | 5’-FAM-AG GTC CCT C/ZEN/T GGC GCT TGT GGC GAG-IABkFQ-3’ | 250 nM | |

Table S2 | Summary statistics for each parameter measured during 24-hour primary influent sampling at WWTP A and B. Summary statistics are also shown for various transformations of the primary influent SARS-CoV-2 RNA counts.

| | Flow (MGD) | BRSV Recovery (%) | PMMoV RNA (CN per L) | SARS-CoV-2 RNA (CN per L) | Recovery Adjusted SARS-CoV-2 RNA (N1 CN per L) | SARS-CoV-2 RNA Load (N1 CN per day) | Recovery Adjusted SARS-CoV-2 RNA Load (N1 CN per day) | SARS-CoV-2 RNA/PMMoV RNA Ratio (log10 N1 CN per L / log10 CN per L) |
|---|---|---|---|---|---|---|---|---|
| WWTP A 6-18&19 | | | | | | | | |
| $\bar{x}$ | 15.1 | 1.89 | $7.64x10^{6}$ | $1.21x10^{3}$ | $3.08x10^{5}$ | $7.04x10^{10}$ | $1.84x10^{13}$ | 0.430 |
| s | 1.55 | 3.90 | $5.16x10^{6}$ | $1.22x10^{3}$ | $3.42x10^{5}$ | $7.35x10^{10}$ | $2.09x10^{13}$ | 0.057 |
| 25th | 14.8 | 0.200 | $3.95x10^{6}$ | 509 | $3.69x10^{4}$ | $2.96x10^{10}$ | $2.09x10^{12}$ | 0.383 |
| 50th | 15.7 | 0.560 | $6.45x10^{6}$ | 667 | $1.49x10^{5}$ | $3.69x10^{10}$ | $9.19x10^{12}$ | 0.418 |
| 75th | 16.2 | 1.70 | $8.78x10^{6}$ | 1490 | $5.36x10^{5}$ | $8.35x10^{10}$ | $3.24x10^{13}$ | 0.452 |
| CoV (%) | 10.2 | 206 | 67.5 | 100 | 111 | 104 | 113 | 13.3 |
| WWTP A 12-2 | | | | | | | | |
| $\bar{x}$ | 14.8 | 0.913 | $7.91x10^{6}$ | $5.82x10^{3}$ | $3.13x10^{6}$ | $3.19x10^{11}$ | $1.75x10^{14}$ | 0.536 |
| s | 1.18 | 1.49 | $3.59x10^{6}$ | $4.08x10^{3}$ | $2.85x10^{6}$ | $2.05x10^{11}$ | $1.50x10^{14}$ | 0.041 |
| 25th | 13.8 | 0.075 | $6.22x10^{6}$ | $2.95x10^{3}$ | $7.00x10^{5}$ | $1.67x10^{11}$ | $4.14x10^{13}$ | 0.504 |
| 50th | 15.2 | 0.15 | $6.77x10^{6}$ | $4.47x10^{3}$ | $2.84x10^{6}$ | $2.51x10^{11}$ | $1.60x10^{14}$ | 0.530 |
| 75th | 15.9 | 1.40 | $10.18x10^{6}$ | $8.70x10^{3}$ | $4.94x10^{6}$ | $4.76x10^{11}$ | $2.99x10^{14}$ | 0.576 |
| CoV (%) | 7.97 | 164 | 45.4 | 70 | 90.8 | 64.3 | 85.7 | 7.70 |
| WWTP B 5-7&8** | | | | | | | | |
| $\bar{x}$ | 8.46 | 0.332 | $1.13x10^{7}$ | 213 | $8.56x10^{4}$ | $6.87x10^{9}$ | $2.72x10^{12}$ | 0.326 |
| s | 1.16 | 0.169 | $3.95x10^{6}$ | 34.6 | $6.49x10^{4}$ | $1.11x10^{9}$ | $1.85x10^{12}$ | 0.013 |
| 25th | 7.68 | 0.185 | $7.10x10^{6}$ | 184 | $4.41x10^{4}$ | $6.06x10^{9}$ | $1.46x10^{12}$ | 0.316 |
| 50th | 8.35 | 0.345 | $1.21x10^{7}$ | 216 | $6.26x10^{4}$ | $6.48x10^{9}$ | $1.99x10^{12}$ | 0.326 |
| 75th | 9.47 | 0.493 | $1.47x10^{7}$ | 244 | $1.20x10^{5}$ | $8.20x10^{9}$ | $4.14x10^{12}$ | 0.338 |
| CoV (%) | 13.7 | 50.9 | 34.9 | 16.2 | 75.9 | 16.3 | 68.1 | 3.96 |
| WWTP B 12-1&2 | | | | | | | | |
| $\bar{x}$ | 7.98 | 0.224 | $4.29x10^{6}$ | $1.51x10^{3}$ | $1.65x10^{6}$ | $4.58x10^{10}$ | $5.37x10^{13}$ | 0.465 |
| s | 1.37 | 0.274 | $1.52x10^{6}$ | $1.07x10^{3}$ | $1.66x10^{6}$ | $3.50x10^{10}$ | $5.62x10^{13}$ | 0.035 |
| 25th | 6.55 | 0.060 | $2.71x10^{6}$ | 720 | $3.73x10^{5}$ | $2.17x10^{10}$ | $1.09x10^{13}$ | 0.431 |
| 50th | 8.65 | 0.100 | $4.53x10^{6}$ | $1.28x10^{3}$ | $1.16x10^{6}$ | $3.23x10^{10}$ | $3.40x10^{13}$ | 0.459 |
| 75th | 9.01 | 0.360 | $5.29x10^{6}$ | $2.08x10^{3}$ | $2.69x10^{6}$ | $5.73x10^{10}$ | $9.13x10^{13}$ | 0.500 |
| CoV (%) | 17.1 | 122 | 35.5 | 70.5 | 100 | 76.4 | 105 | 7.53 |

**SARS-CoV-2 RNA was only detected in 6 of 12 grab samples; non-detects were excluded from summary statistics calculati

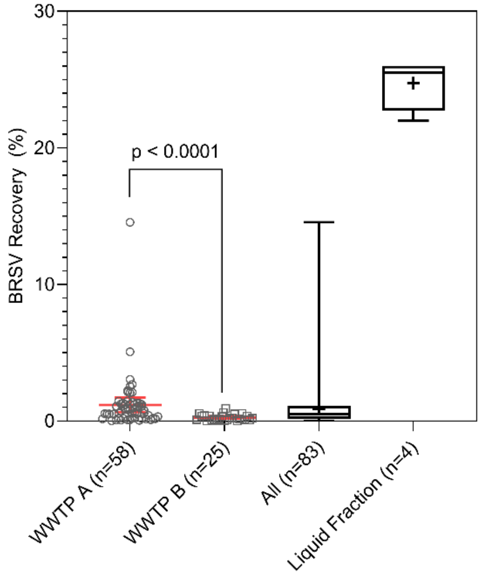


Figure S1 | Process control (BRSV) recovery efficiency for primary influent samples concentrated by adsorption direct-extraction from WWTP A (individual data points, mean and standard deviation), WWTP B (same), and pooled for all primary influent samples. Recovery efficiency for a subset of four primary influent samples concentrated after the suspended solids were removed by centrifugation is also shown. Box plots display minimum, maximum, interquartile range, median, and mean (+). The recovery efficiency from WWTP A was greater than WWTP B ($p < 0.0001$).

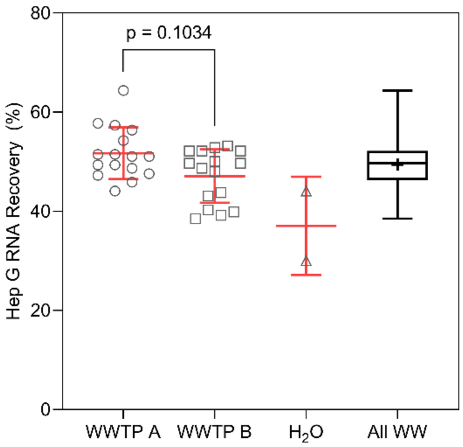


Figure S2 | Recovery efficiency of the HepG extraction and molecular control for primary influent samples concentrated by adsorption direct-extraction from WWTP A (n=16), WWTP B (n=16), PCR-grade water (n=2), and a pooled boxplot of all primary influent samples. Plots for each WWTP and for $H_2O$ display individual data points, mean, and standard deviation. The box plot displays the minimum, maximum, interquartile range, median, and mean (+). There was no difference in the recovery efficiency of the HepG control between primary influent extracts from WWTP A and B ($p = 0.1034$).

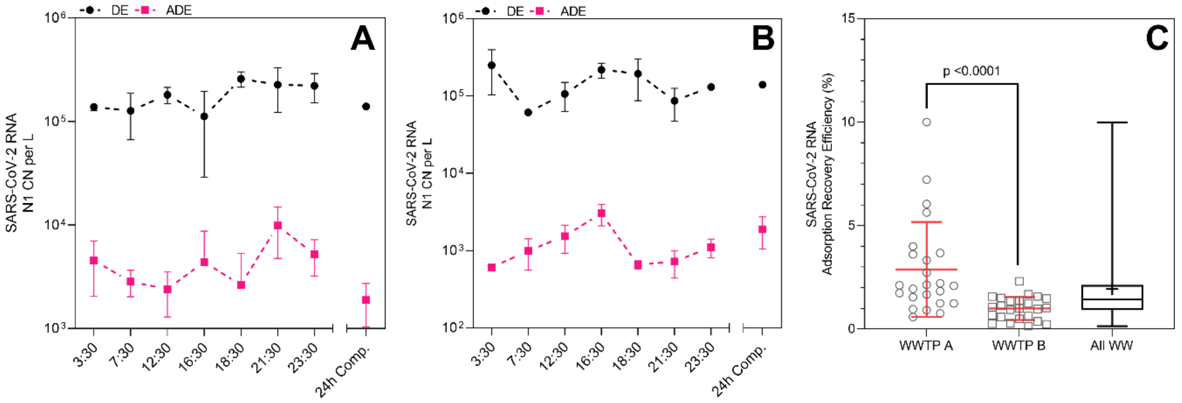


Figure S3 | The mean and standard deviation of SARS-CoV-2 RNA N1 copy number (CN) per liter as measured via direct extraction (DE) of primary influent versus adsorption concentration followed by extraction (ADE) at WWTP A (A) and WWTP B (B) during the 24-hour sampling in December 2020. The SARS-CoV-2 RNA adsorption recovery efficiency was estimated for each WWTP (C) with individual RT-ddPCR data points, mean, and standard deviation at each WWTP shown and a boxplot displaying the minimum, maximum, interquartile range, median, and mean (+) across all RT-ddPCR replicates. SARS-CoV-2 RNA concentration recovery efficiency was significantly greater at WWTP A than WWTP B ($p < 0.0001$).

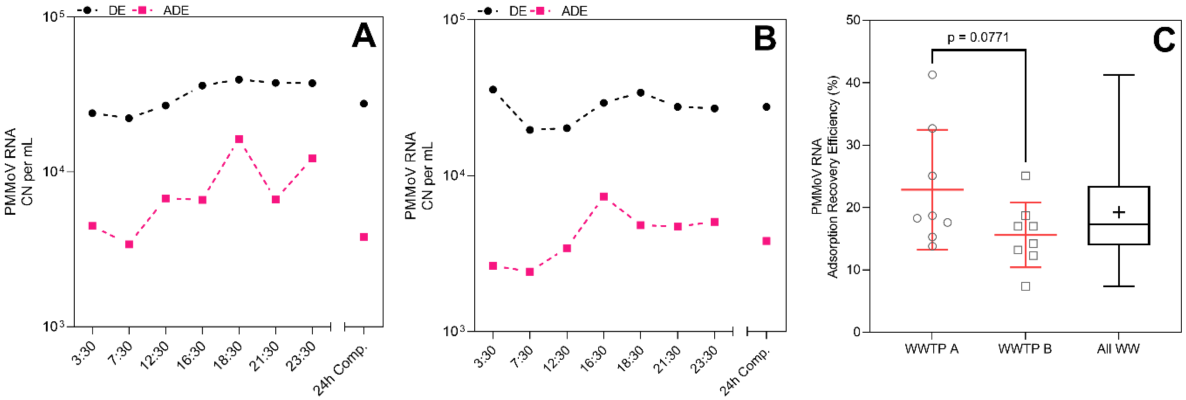


Figure S4 | Mean PMMoV RNA copy number (CN) per milliliter as measured via direct extraction (DE) of influent versus adsorption concentration followed by direct extraction (ADE) at WWTP A (A) and WWTP B (B) during the 24-hour influent sampling in December 2020. The mean and standard deviation of PMMoV RNA adsorption recovery was estimated for each WWTP (C) with individual RT-ddPCR data points, mean, and standard deviation at each WWTP shown and a boxplot displaying the minimum, maximum, interquartile range, median, and mean (+) across all RT-ddPCR replicates. PMMoV RNA concentration recovery efficiency was similar ($p = 0.0771$) between the two WWTPs.

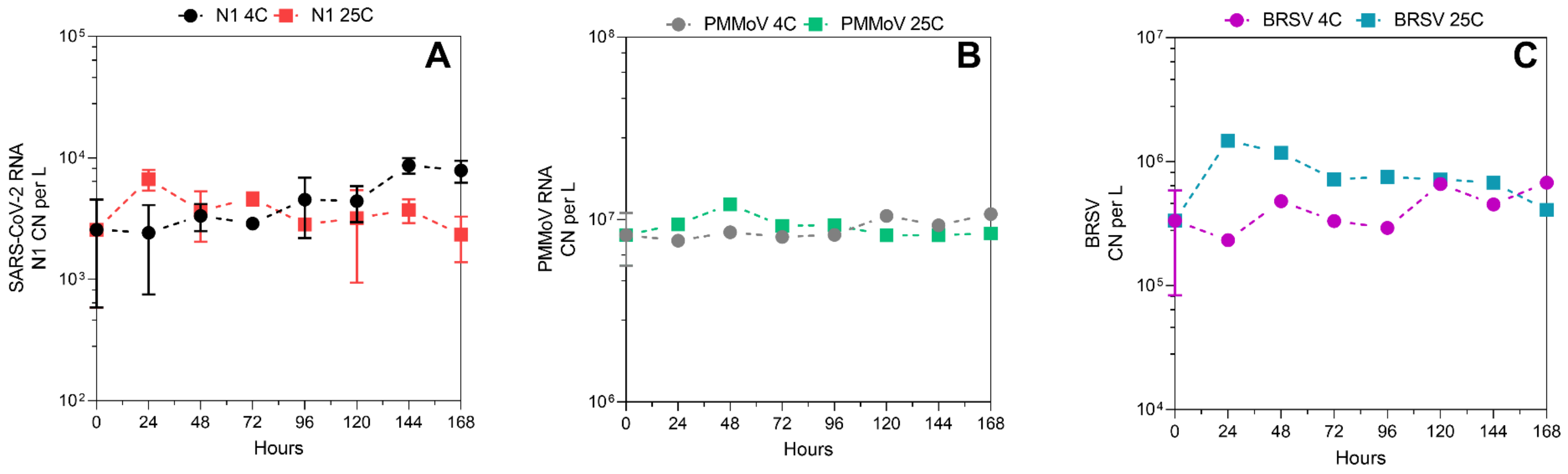


Figure S5 | SARS-CoV-2 RNA N1 copy number (CN) per liter (A), PMMoV RNA CN per liter (B), and BRSV RNA CN per liter (C) enumerated every 24 hours over 7 days (168 hours) of incubation at 4°C and 25°C in primary influent from WWTP A. Plots display mean and standard deviation (where possible).

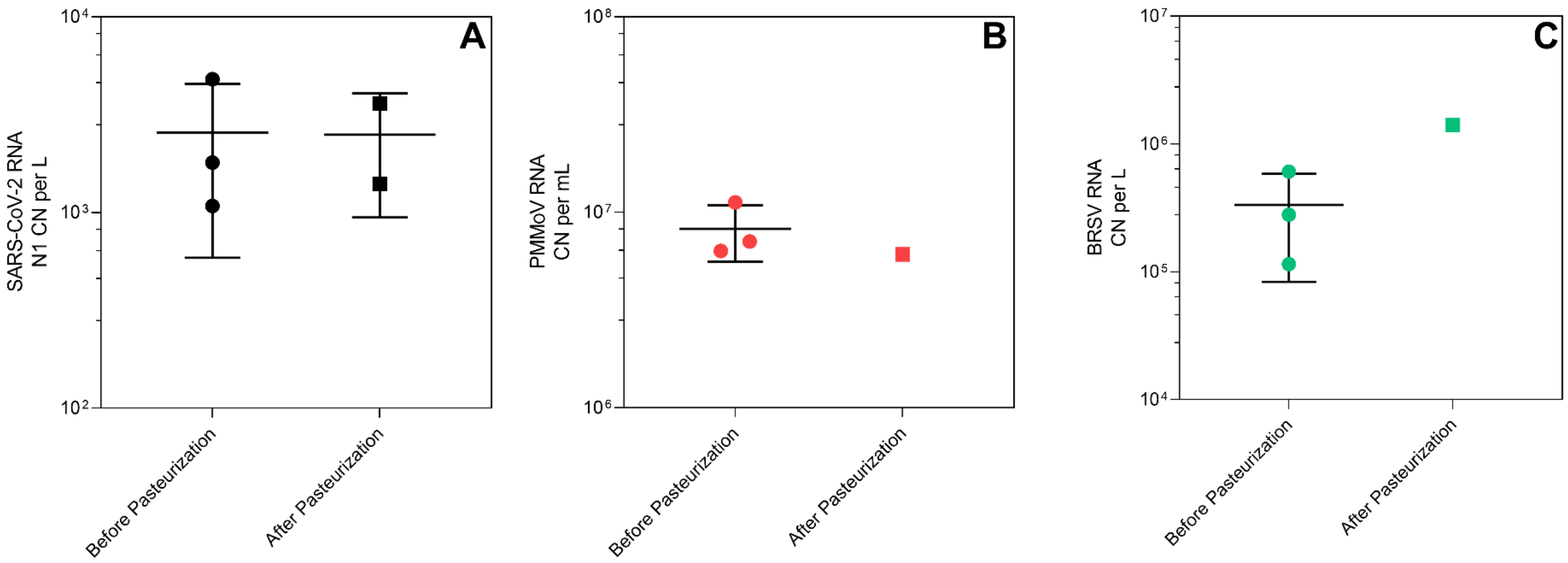


Figure S6 | Measurements of SARS-CoV-2 RNA N1 CN per liter (A), PMMoV RNA CN per liter (B), and BRSV RNA CN per liter (C) before and after pasteurization for 90 minutes at 60°C in primary influent from WWTP A. Plots display individual replicate measurements, mean, and standard deviation (where available).

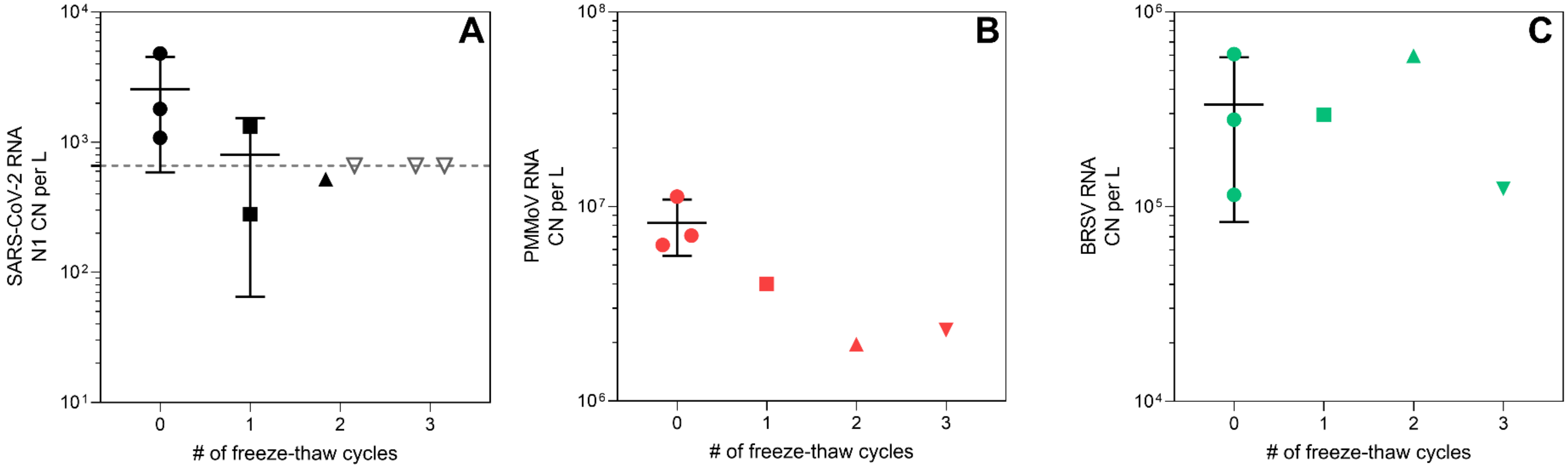


Figure S7 | Measurements of SARS-CoV-2 RNA N1 CN per liter (A), PMMoV RNA CN per liter (B), and BRSV RNA CN per liter (C) following sequential freeze-thaw cycles of primary influent from WWTP A. Plots display individual replicate measurements, mean, and standard deviation (where available).

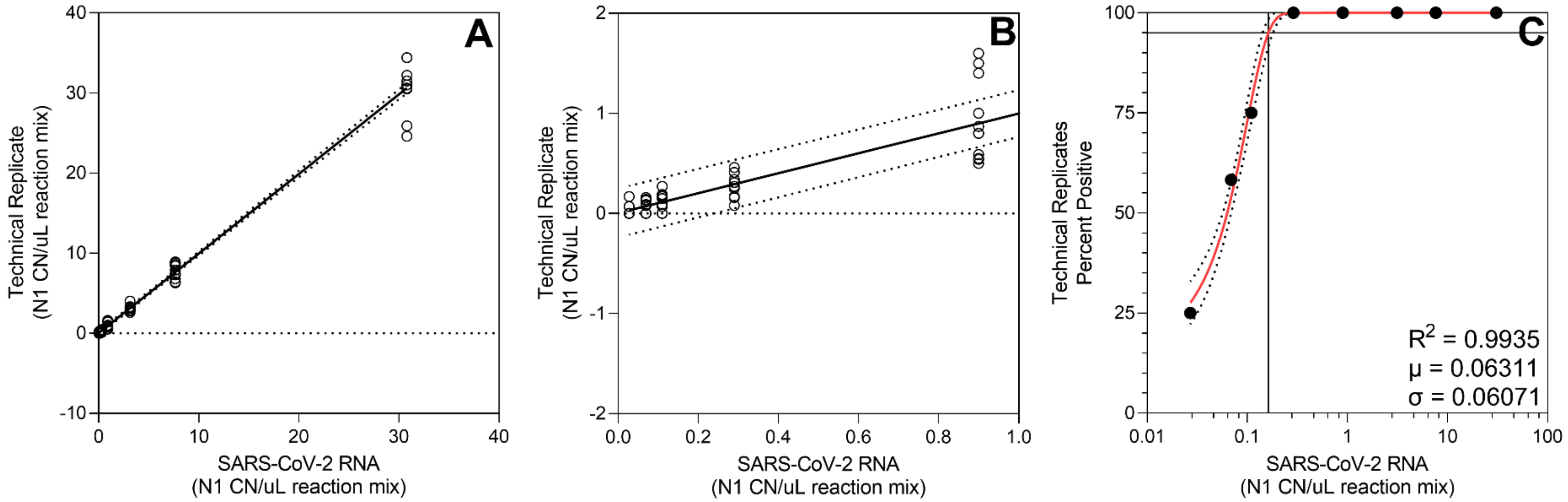


Figure S8 | Quantification of SARS-CoV-2 RNA N1 copy numbers in RT-ddPCR technical replicates along a 1:3 dilution series of RNA positive control material (A,B). A Gaussian distribution fit to the proportion of positive technical replicates along the dilution series (C) and used to estimate the N1 95% LOD of 3.3 copies (95% CI: 2.8 – 3.8) per RT-ddPCR reaction.

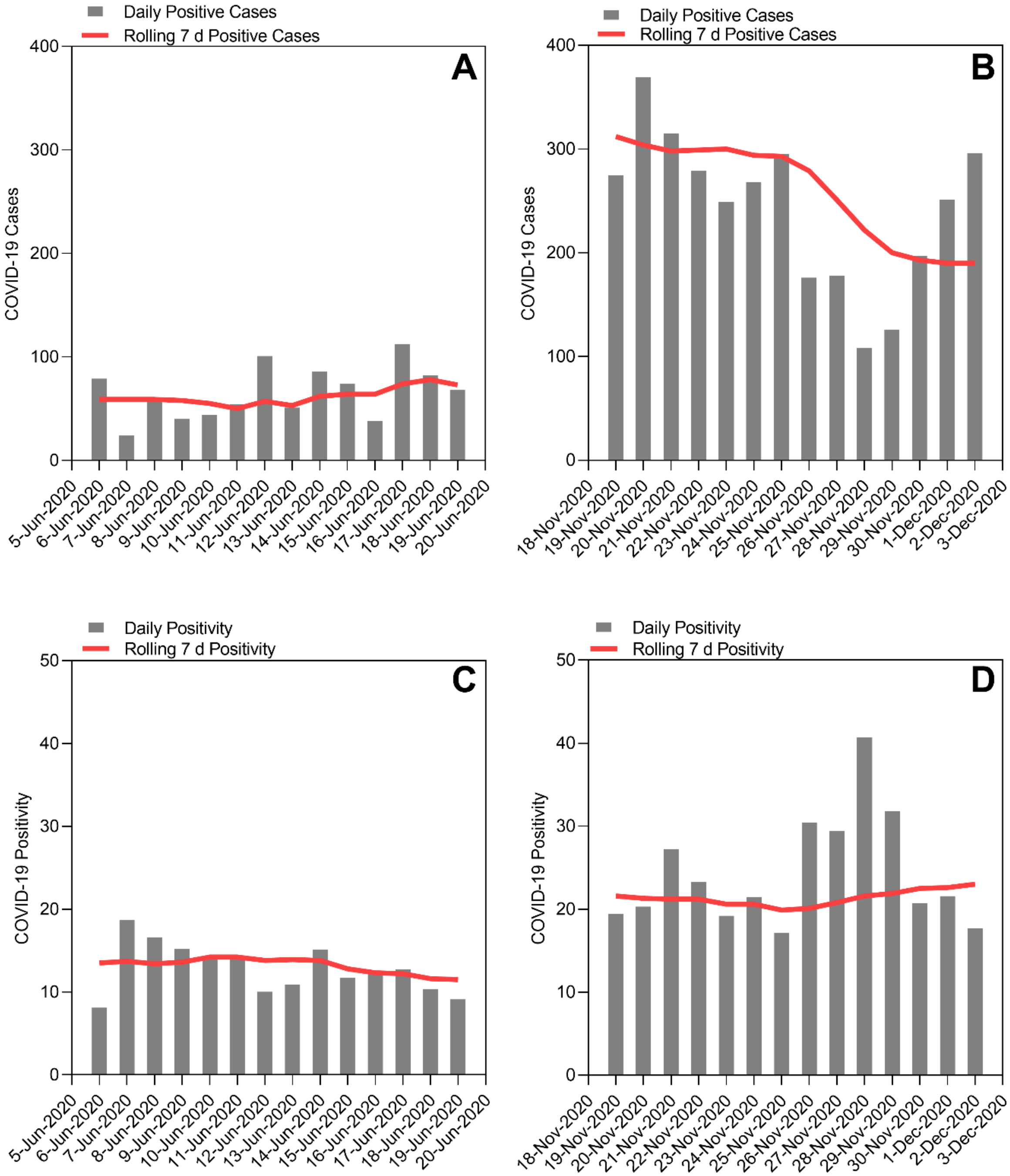


Figure S9 | Daily new cases, positivity, and their associated 7-day rolling averages for COVID-19 in the county containing WWTP A during the two weeks immediately prior to 24-hour influent sampling on June 18 and 19, 2020 (A & C) and December 2, 2020 (B & D).

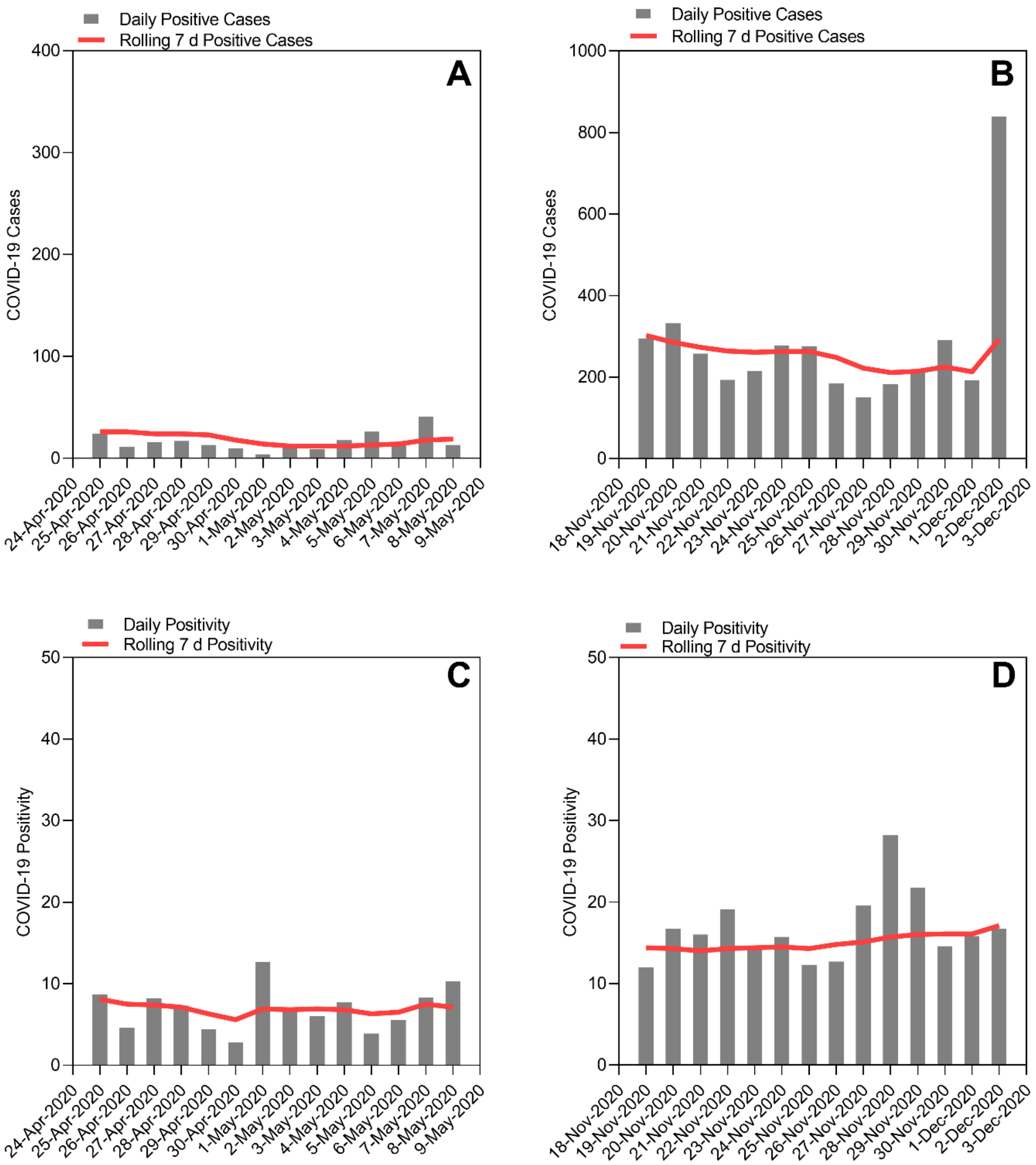


Figure S10 | Daily new cases, positivity, and their associated 7-day rolling averages for COVID-19 in the county containing WWTP B during the two weeks immediately prior to 24-hour influent sampling on May 8 and 19, 2020 (A & C) and December 1 and 2, 2020 (B & D).

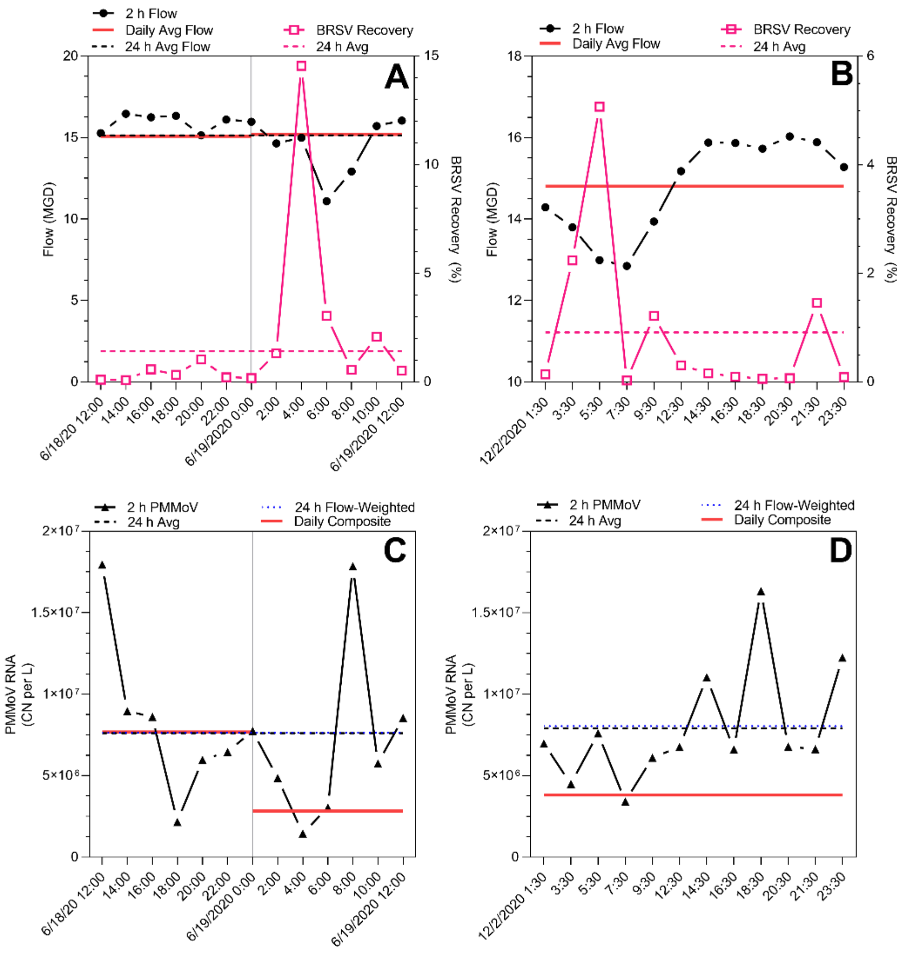


Figure S11 | WWTP A primary influent characteristics as observed during two 24-hour sampling events: influent flow rates and process control recovery efficiency (A & B), and PMMoV RNA copy number density (C & D). Data displayed include the observations in serial grab samples, the average observed over the 24-hour interval, the flow-weighted 24-hour average, and the daily composite sample observation (when available).

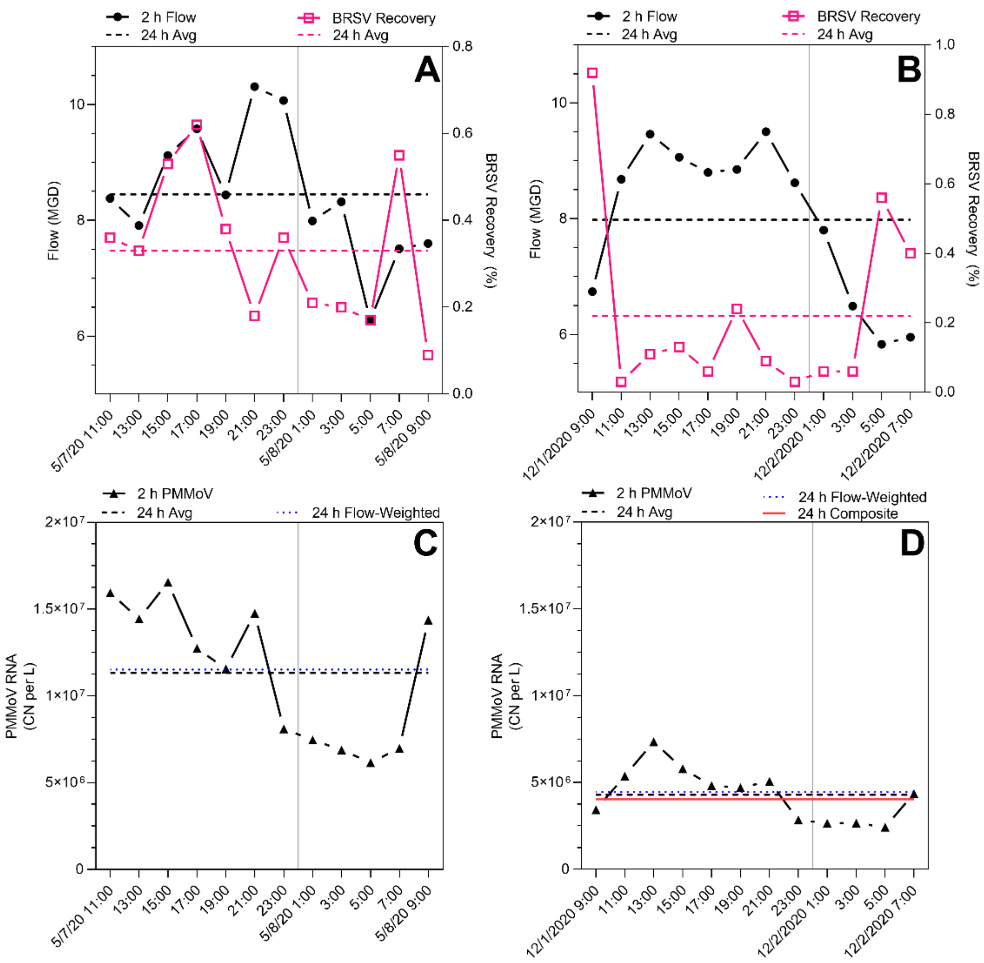


Figure S12 | WWTP B primary influent characteristics as observed during two 24-hour sampling events: influent flow rates and process control recovery efficiency (A & B), and PMMoV RNA copy number density (C & D). Data displayed include the observations in serial grab samples, the average observed over the 24-hour interval, the flow-weighted 24-hour average, and the daily composite sample observation (when available).

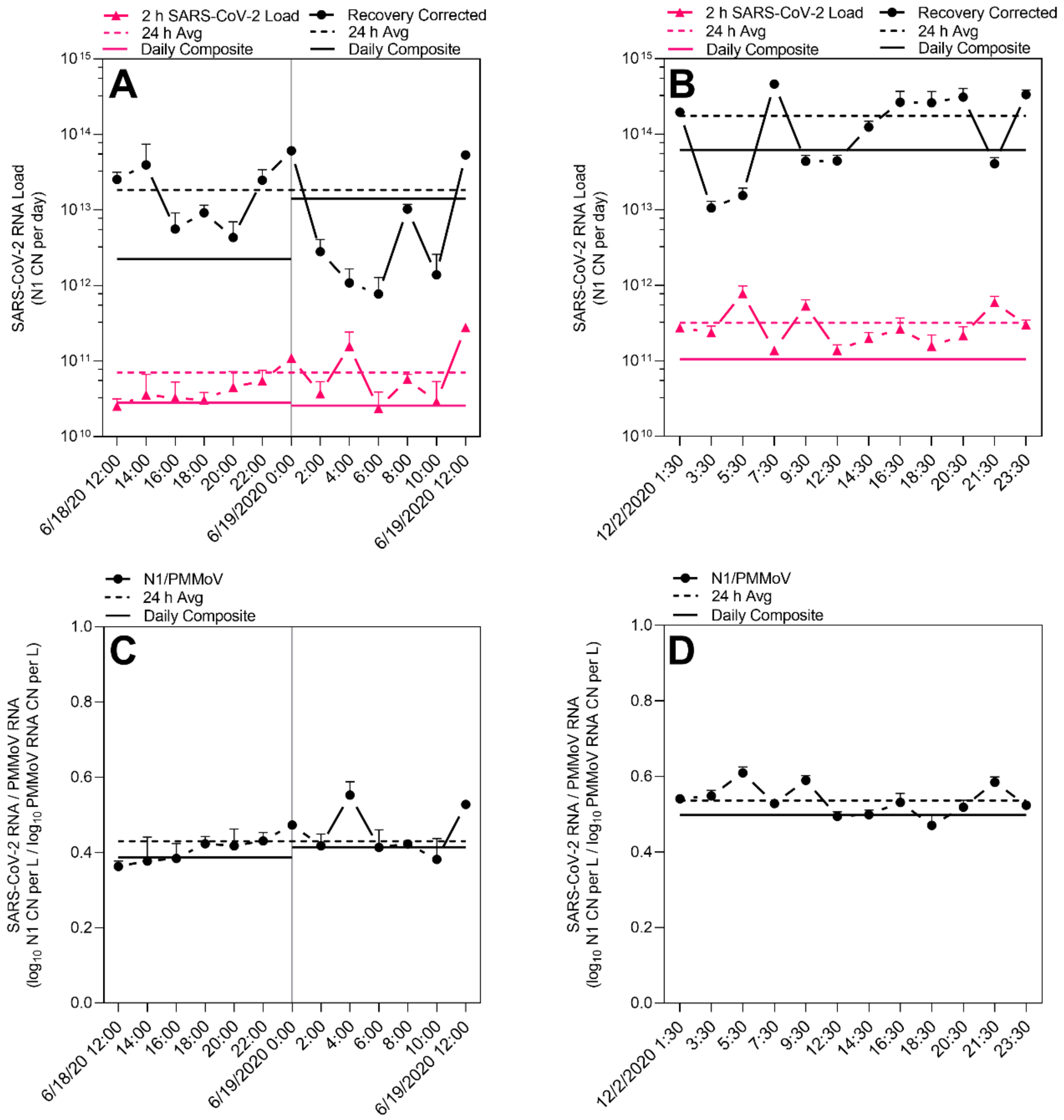


Figure S13 | WWTP A primary influent characteristics as measured during two 24-hour sampling events: influent SARS-CoV-2 loading, copy number (CN) per day (concentration x flow), with and without recovery adjustment (A & B), and the ratio of the $log_{10}$ SARS-CoV-2 RNA copy number per liter to $log_{10}$ PMMoV RNA copy number per liter (C & D). Data displayed include the observations in serial grab samples, the average observed over the 24-hour interval, and the daily composite sample observation (when available).

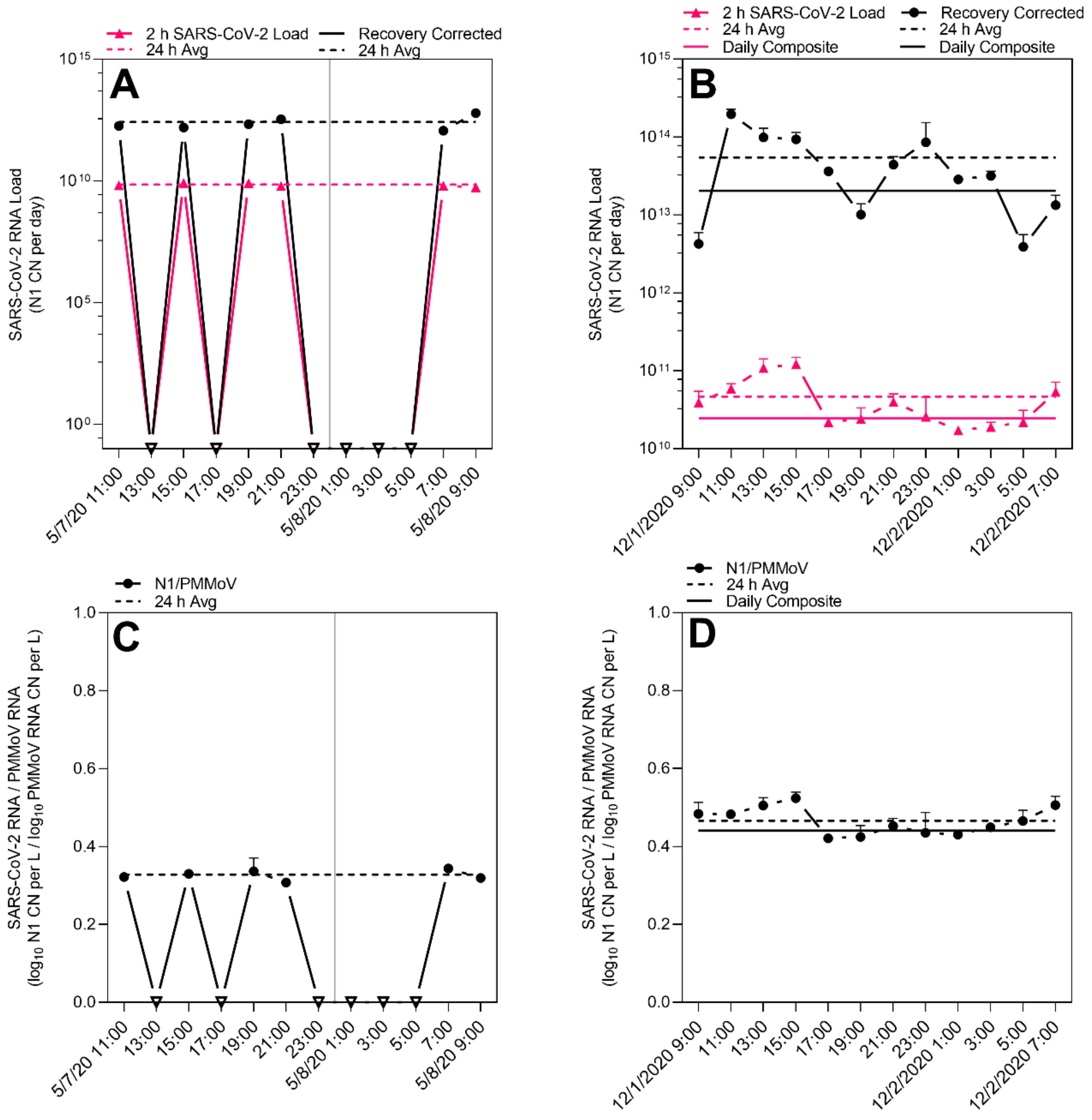


Figure S14 | WWTP B primary influent characteristics as measured during two 24-hour sampling events: influent SARS-CoV-2 loading, copy number (CN) per day (concentration x flow), with and without recovery adjustment (A & B), and the ratio of the $\log_{10}$ SARS-CoV-2 RNA copy number per liter to $\log_{10}$ PMMoV RNA copy number per liter (C & D). Data displayed include the observations in serial grab samples, the average observed over the 24-hour interval, and the daily composite sample observation (when available).

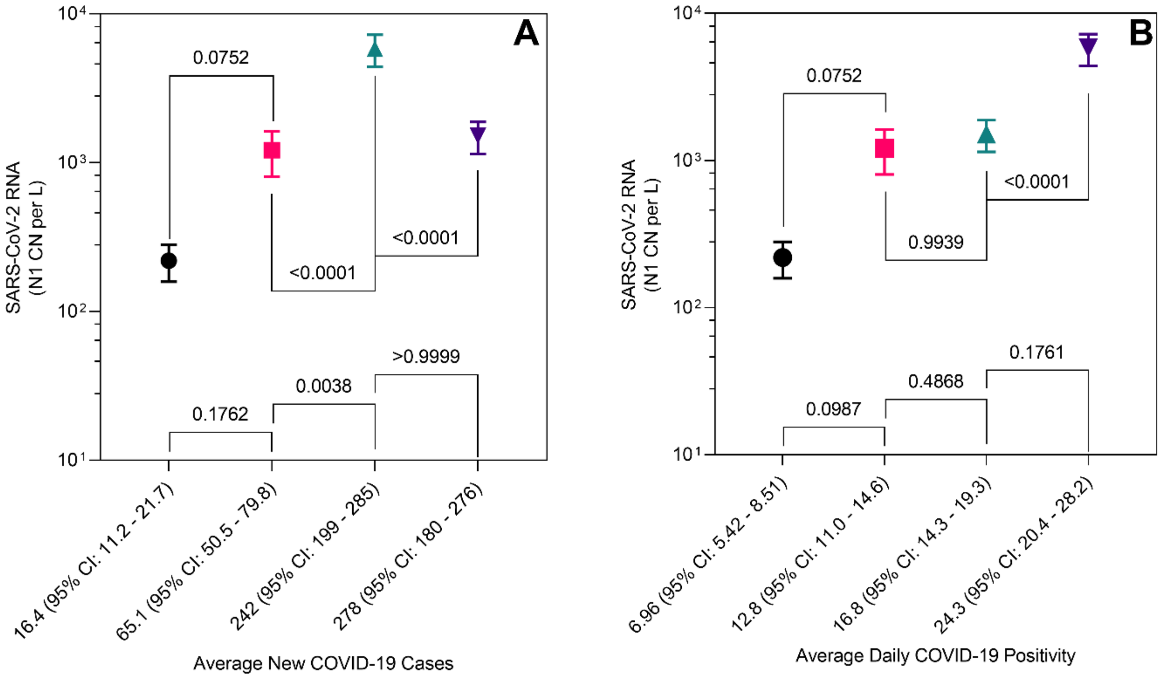


Figure S15 | SARS-CoV-2 RNA N1 concentrations, copy number (CN) per liter, in primary influent stratified by ordinal arrangement of average daily COVID-19 cases (A) and average daily COVID-19 positivity (B). Clinical averages and confidence intervals are calculated for the two weeks prior to the 24-hour primary influent sampling period.
